# Immunological dysfunction persists for 8 months following initial mild-moderate SARS-CoV-2 infection

**DOI:** 10.1101/2021.06.01.21257759

**Authors:** Chansavath Phetsouphanh, David Darley, Daniel B Wilson, Annett Howe, C. Mee Ling Munier, Sheila K Patel, Jennifer A Juno, Louise M Burrell, Stephen J Kent, Gregory J Dore, Anthony D Kelleher, Gail V Matthews

**Author notes:** Correspondence Dr Chansavath Phetsouphanh, Prof. Anthony Kelleher, Prof. Gail Matthews. These authors contributed equally (Chansavath Phetsouphanh, David Darley, Anthony D Kelleher, Gail V Matthews).

## Abstract

A proportion of patients surviving acute COVID-19 infection develop post-COVID syndrome (long COVID) encompassing physical and neuropsychiatric symptoms lasting longer than 12 weeks. Here we studied a prospective cohort of individuals with long COVID compared to age/gender matched subjects without long COVID (from the ADAPT study), healthy donors and individuals infected with other non-SARS CoV2 human coronaviruses (the ADAPT-C study). We found highly activated innate immune cells and an absence of subsets of un-activated naïve T and B cells in peripheral blood of long COVID subjects, that did not reconstitute over time. These activated myeloid cells may contribute to the elevated levels of type I (IFN-β) and III interferon (IFN-λ1) that remained persistently high in long COVID subjects at 8 months post-infection. We found positive inter-analyte correlations that consisted of 18 inflammatory cytokines in symptomatic long COVID subjects that was not observed in asymptomatic COVID-19 survivors. A linear classification model was used to exhaustively search through all 20475 combinations of the 29 analytes measured, that had the strongest association with long COVID and found that the best 4 analytes were: IL-6, IFN-γ, MCP-1 (CCL2) and VCAM-1. These four inflammatory biomarkers gave an accuracy of 75.9%, and an F1 score of 0.759, and have also previously been associated with acute severe disease. In contrast, plasma ACE2 levels, while elevated in the serum of people previously infected with SARS-CoV-2 were not further elevated in subjects with long COVID symptoms. This work defines immunological parameters associated with long COVID and suggests future opportunities to prevention and treatment.

## Introduction

Coronavirus disease 2019 (COVID-19), the illness caused by severe acute respiratory syndrome coronavirus 2 (SARS-CoV-2) infection is characterised by a wide spectrum of clinical severities in the acute phase, ranging from asymptomatic to severe fatal forms ^1,2^. Immunological disturbance during acute illness has been well described and is likely to both play a role in host defence and be involved in the pathogenesis of severe COVID-19 ^3^. Immunological abnormalities described in the acute phase include significant immune dysregulation with lymphopaenia and elevations in IL-2, IL-6, TGF-β, IL-10, GM-CSF and TNF^3,4^.

A proportion of patients surviving acute COVID-19 develop post-COVID syndrome encompassing various physical and neuropsychiatric symptoms lasting longer than 12 weeks, also known as chronic COVID syndrome (CCS), post-acute sequelae of COVID-19 (PASC) and long-haul COVID (long COVID) ^5-8^. Although well described following other viral infections including Severe Acute Respiratory Syndrome Coronavirus (SARS-CoV-1)^9-11^ and Middle East Respiratory Syndrome Coronavirus (MERS-CoV)^12^, these syndromes appear commonly following SARS-CoV-2 infection including following initial mild-moderate illness ^13-16^. Persistent symptoms 6 months after hospitalization were observed in 76% of discharged patients in a study by Huang *et al*, with the most frequent symptoms reported being muscle weakness and fatigue ^17,18^. Amongst community managed COVID-19 cases, prevalence of persisting symptoms is lower, but remains higher than might be expected given the severity of the initial illness, affecting between 10-30% of individuals at 2-3 months post infection, ^19,20^ continuing out to at least 8 months after initial illness ^21^. The spectrum of post-acute sequelae of COVID-19 (henceforth denoted as long COVID) is broad encompassing as many as 50 plus symptoms although characteristically manifests as severe relapsing fatigue often accompanied by dyspnoea, chest tightness, cough and headache ^22^. The underlying pathophysiology of long COVID is poorly understood and there is currently little evidence defining the mechanisms of systemic inflammation following COVID-19 and any immune correlates of ongoing symptoms.

We examined a prospective cohort of individuals followed systematically post confirmed COVID-19 infection (the ADAPT study) and compared them to healthy donors and individuals contemporaneously infected with other non-SARS CoV2 human coronaviruses (HCoV, ADAPT-C) to firstly characterise their immunological features up to 8 months post COVID-19 infection and secondly to compare those with and without persisting symptoms at this time point. Serum profiles of 29 analytes from 62 patients, 31 with long COVID and 31 matched asymptomatic controls were evaluated at 5- and 8-months post SARS-CoV-2 infection. Immune profiles identified in COVID-19 were compared with subjects who had been infected with common cold coronaviruses (HCoV-NL63, O229E, OC43 or HKU1) and unexposed healthy donors.

## Methods

### Cohort Characteristics

The ADAPT study is a prospective cohort study of post-COVID-19 recovery established in April 2020 ^21^. 147 participants with confirmed SARS-CoV-2 infection were enrolled, the majority following testing in community based clinics run by St Vincent’s Hospital Sydney, with some patients also enrolled with confirmed infection at external sites. Initial study follow-up was planned for 12 months post-COVID-19, and subsequently extended to 2 years. Extensive clinical data and a biorepository was systematically collected prospectively. The aims of ADAPT are to evaluate a number of outcomes after COVID-19 relating to pathophysiology, immunology and clinical sequalae. Laboratory testing for SARS-CoV-2 was performed using nucleic acid detection from respiratory specimens with the EasyScreen™ Respiratory Detection kit (Genetic Signatures, Sydney, Australia), and the EasyScreen™ SARS-CoV-2 Detection kit. Two ADAPT cohort sub-populations were defined based on initial severity of COVID-19 illness; 1. patients managed in the community and 2. patients admitted to hospital for acute infection (including those requiring intensive care support for acute respiratory distress syndrome (ARDS). Patients were defined as ‘long COVID’ at 4-months based on the presence of any ≥ 1 of the following; fatigue, dyspnoea or chest pain ^21^. These patients were gender and age (+/- 10 years) matched with ADAPT participants without long COVID (Matched ADAPT controls) (Table 1). Samples for these analyses were collected at the 3-, 4- and 8-month assessments. Our cohort of n=62 (31 long COVID and 31 matched controls); enrolment visits were performed at median 76 (IQR 64-93) days after initial infection. Their 4 month assessments were performed at median 128 (IQR 115-142) days after initial infection (4.2 months). Their 8 month assessments were performed at median 232 (IQR 226-253) days after initial infection (7.7 months). 4 participants did not complete the 8-month assessment after the 4-month assessment. The reasons for this include-did not attend (n=2) and Lost to Follow-up (n=2). A further population of patients presenting to St Vincent’s Hospital clinics for COVID-19 testing on the multiplex respiratory panel who were PCR positive for any of the 4 human common cold coronaviruses (HCoV-NL63, O229E, OC43 or HKU1) and PCR negative for SARS-CoV-2 were recruited into the ADAPT-C sub study and used as a comparator group.

**Table 1.**
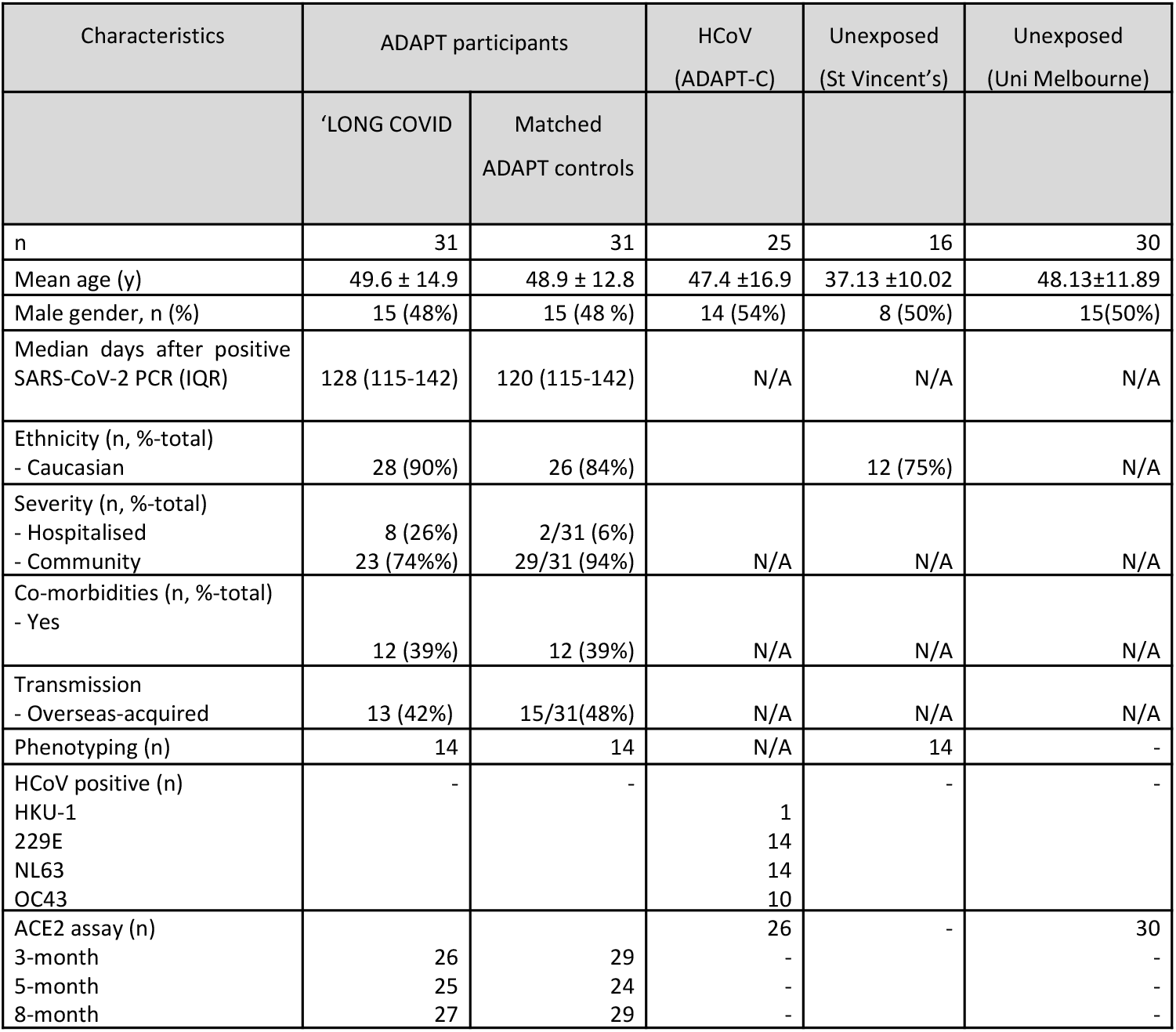
Patient characteristics. Age, gender, ethnicity and co-morbidities within the cohorts sampled. HCoV patients who were PCR positive for 229E were also positive for NL63.

### Ethics

The ADAPT study was approved by the St Vincent’s Hospital Research Ethics Committee (2020/ETH00964) and is a registered trial (ACTRN12620000554965). ADAPT-C sub study was approved by the same committee (2020/ETH01429). All data were stored using REDCap electronic data capture tools. Unexposed healthy donors were recruited through St Vincent’s Hospital and was approved by St Vincent’s Hospital Research Ethics Committee (HREC/13/SVH/145). The University of Melbourne unexposed donors were approved by Medicine and Dentistry HESC-Study ID 2056689. All participants gave written informed consent.

### Sample processing and Flow cytometry

Blood was collected for biomarker analysis (SST 8.5mLs x 1 (Serum) and EDTA 10mLs x 1 (Plasma)) and 36mLs was collected for PBMCs (ACD 9mLs x4). Phenotyping of PBMC was performed as described previously ^23^. Briefly, cryopreserved PBMCs were thawed using RPMI (+L-glut) medium (ThermoFisher Scientific, USA) supplemented with Penicillin/Streptomycin (Sigma-Aldrich, USW), and subsequently stained with antibodies binding to extracellular markers. Extracellular panel included: Live/Dead dye Near InfraRed, CXCR5 (MU5UBEE), CD38 (HIT2) (ThermoFisher Scientific, USA); CD3 (UCHT1), CD8 (HIL-72021), PD-1 (EH12.1), TIM-3 (TD3), CD27 (L128), CD45RA (HI100), IgD (IA6-2), CD25 (2A3), and CD19 (HIB19) (Biolegend, USA); CD4 (OKT4), CD127 (A019D5), HLA-DR (L234), GRP56 (191B8), CCR7 (G043H7) and CD57 (QA17A04) (BD Biosciences, USA). Perm Buffer II (BD Pharmingen) was used for intracellular staining of granzyme B (GB11, BD Biosciences). Samples were acquired on an Cytek Aurora (Biolegend, USA) using the Spectroflo software. Prior to each run, all samples were fixed in 0.5% Paraformaldehyde.

### Serum Analytes

LEGENDplex^™^ Human Anti-Virus Response Panel (IL-1β, IL-6, IL-8, IL-10, IL-12p70, IFN-α2, IFN-β, IFN-λ1, IFN-λ2/3, IFN-γ, TNF-α, IP-10, GM-CSF) and custom-made panel (IL-5, IL-9, IL-13, IL-33, PD-1, sTIM-3, sCD25, CCL2(MCP-1), Pentraxin-3(PTX3), TGF-β1, CXCL9(MIG-1), MPO, PECAM-1, ICAM-1, VCAM-1) were purchased from Biolegend (San Diego, CA, USA) and assays performed as per manufacturer’s instructions. Beads were acquired and analysed on BD Fortessa X20 SORP (BD Biosciences, San Jose, CA, USA). Samples were run in duplicate, and 4000 beads were acquired per sample. Data analysis was performed using Qognit LEGENDplex^™^ software (Biolegend, San Diego, CA, USA). Lower limit of detection (LOD) was values were used for all analytes at the lower limit.

### Catalytic ACE2 detection in plasma

Plasma ACE2 activity was measured using a validated, sensitive quenched fluorescent substrate-based assay as previously described ^24^. Briefly, plasma (0.25 ml) was diluted into low-ionic-strength buffer (20 mmol/L Tris-HCl, pH 6.5) and added to 200 *m*l ANXSepharose 4 Fast-Flow resin (Amersham Biosciences, GE Healthcare, Uppsala, Sweden) that removed a previously characterized endogenous inhibitor of ACE2 activity. After binding and washing, the resulting eluate was assayed for ACE2 catalytic activity. Duplicate samples were incubated with the ACE2-specific quenched fluorescent substrate, with or without 100 mM ethylenediaminetetraacetic acid. The rate of substrate cleavage was determined by comparison to a standard curve of the free fluorophore, 4-amino-methoxycoumarin (MCA; Sigma, MO, USA) and expressed as *r*mole of substrate cleaved/mL of plasma/min. The intra-assay and inter-assay coefficient of variation was 5.6% and 11.8% respectively. Samples below the limit of detection were designated 0.02 (half the LOD – ie 50% x 0.04).

### Linear Model

The four analytes most associated with long COVID were identified via Linear Classification. For an arbitrary set of four analytes, let the concentration of the i^th^ analyte after four and eight months be denoted 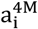 and 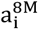, respectively. Linear Classification assigns two weights 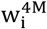 and 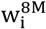 to each analyte. A linear function of these concentrations and weights takes the form 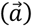 is a threshold parameter. The weights 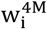 and 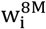 as well as the threshold parameter θ are selected to maximise the predictive power of the linear classifier by training on the analyte data.

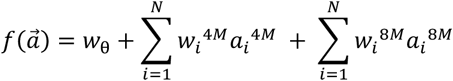

The data are standardized prior to training, each analyte was rescaled to have zero mean and unit variance across all participants, thus removing the possibility of a single analyte dominating the classifier. Then for a given set of four analytes we stratified our data into a training set and a testing set. Due to the moderately small sample size of 58 participants we choose our training set to include all but one participant. The excluded participant does not influence the training of the classifier and is used to test the accuracy of the classifier. There are four possible outcomes; true positive (TP: both the classifier and data say the participant had long COVID), true negative (TN: both the classifier and data say the participant had asymptomatic COVID), false positive (FP: classifier predicts the participant will have long COVID but the data disagrees) and false negative (FN: classifier predicts the participant will have asymptomatic COVID but the data disagrees). Each participant is removed in turn to be the test data, and thus a total of 58 classifiers were trained and the number of TPs, TNs, FPs and FNs were recorded. This step is known as cross validation and removes the possibility of bias in the way the training and test data is selected. Thus for a given set of four analytes we measure how well they predict long COVID with two metrics.

Firstly, the accuracy is defined as TP+TN/𝓃 and measures the proportion of test participants that had their COVID status correctly predicted. The second measure is the F1 score and is defined as TP/(TP + 0.5 × (FP + FN)), which is a measure that combines recall, how many long COVID cases were correctly predicted, and precision, of all the participants predicted to have long COVID how many were correct. The accuracy and F1 score are used to select which combination of four analytes are most associated with long COVID. For four analytes there are a total of 20475 combinations. The best four analytes are selected by identifying the four analytes that achieve the highest accuracy. If two or more sets of four analytes have the highest accuracy we defer to the F1 score to identify the best set of four analytes. The training of the many classifiers was performed in Python3 using the Scikit-learn machine learning package.

### Dimensional Reduction and Clustering Analysis

FCS3.0 files were compensated manually using acquisition-defined matrix as a guide, and gating strategy was based on unstained or endogenous controls. Live singlets were gated from long COVID and asymptomatic matched controls using Flowjo v.10.7.2, samples were decoded and statistical analysis between groups and unsupervised analysis was performed, with matched asymptomatic controls as the primary comparator group. For unsupervised analysis, the following FlowJo plugins were used: DownSample (v.3), TriMap (v.0.2), Phenograph (v.3.0) and ClusterExplorer (v.1.5.9) (all FlowJo LLC). First, 100 000 events per sample were downsampled from the total live singlet gate (Supp. Fig. 1). The newly generated FCS files were labelled according to control or patient group (long COVID or matched controls) and concatenated per group. Subsequently, 20, 000 events were taken from each grouped sample by downsampling. The two new FCS files corresponding to long COVID and matched controls were then concatenated for dimensionality reduction analysis using TriMap (40, 000 events in total). TriMap was conducted using the following parameters-to include the markers CD25, CD38, CCR7, CD19, IgD, CD45RA, PD-1, TIM-3, CD4, CD57, CD127, CD27, HLA-DR, CD8, CXCR5,GPR56 and Granzyme B, and using the following conditions: metric = euclidean, nearest neighbours = 15, and minimum distance = 0.5. Phenograph plugin was then used to determine clusters of phenotypically related cells. The same markers as TriMap and parameters k = 152 and Run ID = auto was used for analysis. Finally, ClusterExplorer plugin was used to identify the phenotype of the clusters generated by phenograph.

### Statistical Analysis

All column graphs are presented as medians with inter-quartile ranges. One-way ANOVA utilising Kruskal-Wallis and Dunn’s correction for multiple comparisons was used for serum analyte analysis. Wilcoxon paired *t* test was used to analyse statistical data employing Prism 9.0 (GraphicPad, La Jolla, CA, USA) software. For unpaired samples Mann-Whitney U test was used. RStudio version 1.2.1335 was used to generate correlograms utilizing corrplot package with Spearman’s method used to determine Rho values and cor.mtest with 95% confidence level was used to determined *p* values. *p* values <0.05 were considered significant (*<0.05, **<0.01, ***<0.001, and ****<0.0001).

## Results

### The ADAPT cohort

All participants were drawn from a single prospective observational cohort designed to follow patients after acute COVID-19 infection. The ADAPT study^16^ enrolled all adults with SARS-CoV-2 infections confirmed by polymerase chain reaction (PCR) at St Vincent’s Hospital community based testing clinics in Sydney (Australia), who were able to be contacted and agreed to participate (70.5% of those diagnosed). Participants were prospectively systematically assessed according to a pre-defined schedule which included routine visits at enrolment, month 4 and month 8. The study was commenced as soon as ethics approval was obtained and for the majority of participants in the first wave, the first enrolment visit occurred somewhere between month 2-3 post infection (median 79 days)^16,21^ with 93.6% and 84.5% of participants completing subsequent month 4 (median 128 days) and month 8 (median 232 days) visits timed from the date of diagnosis of their initial infection (Table 1). The drop-out rate has been very low to date: approximately 9.4% at 12 months. Participants were designated as Long COVID (LC) based on the presence of one of three major symptoms (fatigue, dyspnoea or chest pain) at the month 4 timepoint. These participants were age and gender matched with 31 participants from the same cohort who reported no ongoing symptoms at the 4 month timepoint (designated as matched controls (MC)) (Suppl. Table 1).

The matched control group were symptomatic at acute initial infection (Suppl. Table 2), but did not develop long COVID and were asymptomatic at month 4. In terms of clinical recovery, there was a 10% trend towards some improvement over time, but this was not statistically significant, Fischer Exact p=0.44. With regard to the external enrolments (29.5%), we also allowed enrolment in the study for some participants who were diagnosed outside of the St Vincent’s testing clinic. In these cases participants were required to have documented evidence of a positive swab and entered into the same schedule of assessments for follow-up.

### Elevated levels of pro-inflammatory cytokines following SARS-CoV-2 infection that are maintained at 8 months

In order to assess biomarkers associated with recovery from symptomatic SARS-CoV-2 infection 28 analytes were analysed from serum of convalescent subjects with long COVID (LC) or asymptomatic matched controls (MC) at 4 months post-infection. Individuals that were PCR positive for prevalent common cold coronaviruses (HCoV) and sampled within the same time frame following infection (3-6months) and unexposed healthy donors sampled prior to December 2019, were used as comparator groups. Six pro-inflammatory cytokines were highly elevated in the two COVID-19 convalescent ADAPT groups compared to control groups (Fig. 1A), while no difference was observed with the 22 other analytes (Supp. Fig. 2A&B). It is noteworthy that no difference was observed between long COVID and asymptomatic matched controls for any of the individual analytes examined at 4 months post infection. Type I interferon-β (IFN-β) was 7.92-fold (median values shown in Supp. Fig 2C) and 7.39-fold higher in LC and MC compared to HCoV (both *p<0*.*0001*), and 7.32- and 6.83-fold higher compared to unexposed healthy controls (both *p<0*.*0001*, respectively). Type III interferon-lamdba1 (IFN-λ1) was 2.44-fold and 3.24-fold higher in LC and MC compared to HCoV (both *p<0*.*0001*), and 2.42- and 3.21-fold higher compared to unexposed, (*p<0*.*01 and p<0*.*001*, respectively). This was also the case for IL-8 (CXCL8) levels; LC (3.43-fold) and MC (3.56-fold) higher compared to both HCoV and unexposed (all *p<0*.*0001*). Interferon gamma induced chemokine CXCL10 (IP-10) was elevated in LC and compared to HCoV 2.15-fold (*p<0*.*001*) and unexposed 3.20-fold (*p<0*.*001*), similarly MC group were also significantly higher than comparator groups (1.7-fold, *p<0*.*05* and 3.06-fold, *p<0*.*01*, respectively). CXCL9(MIG-1) was 1.69-fold higher than unexposed (*p<0*.05), and soluble TIM-3 levels were only elevated in LC and not MC when compared to HCoV comparator group 1.46-fold (*p<0*.*05*).

**Figure 1.**
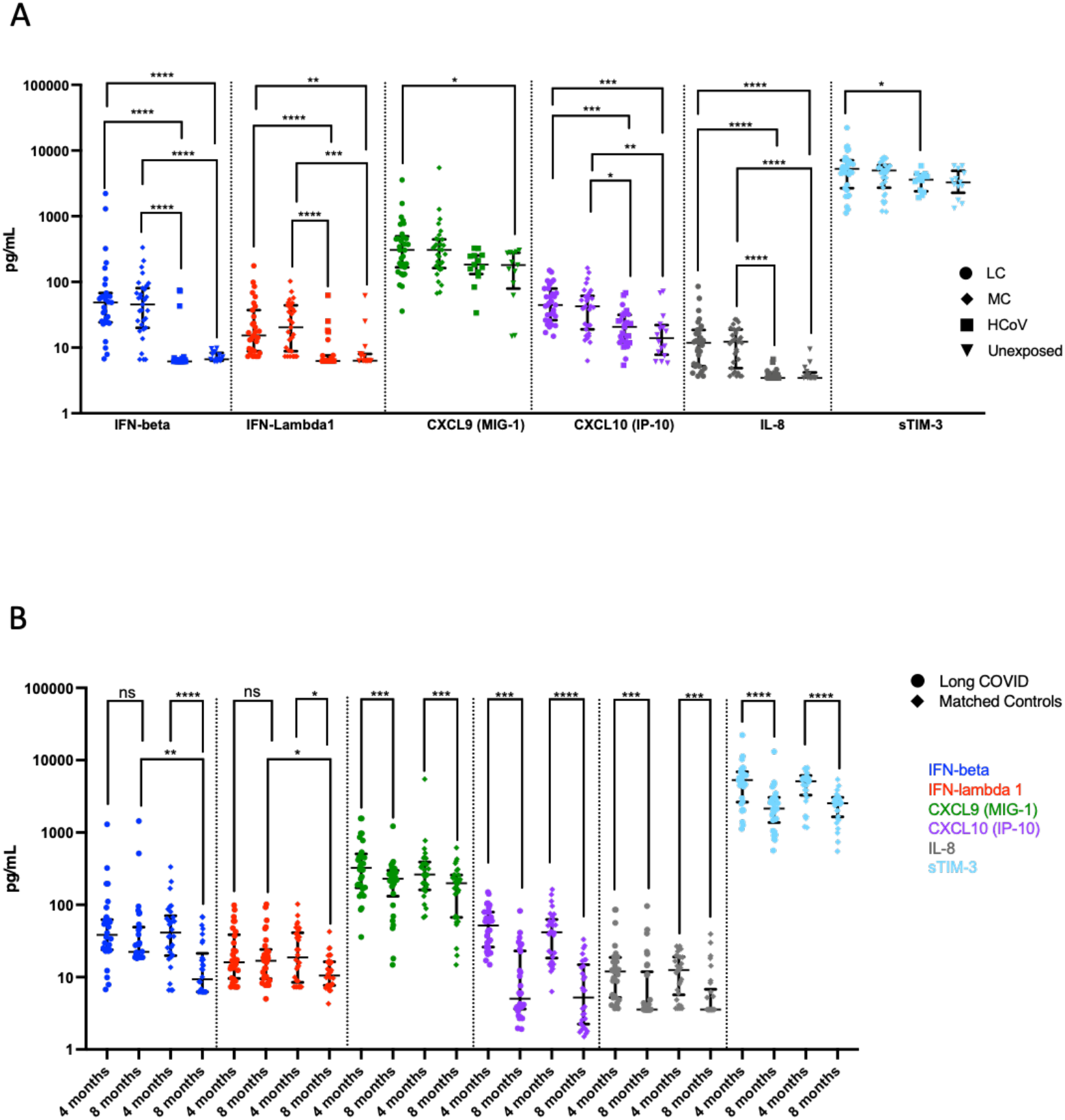
Elevated levels of pro-inflammatory cytokines that persists after month 8 of convalescence. A) Higher levels of IFN-β, IFN-λ1, CXCL9 (MIG-1), CXCL10 (IP-10), IL-8 and sTIM-3 at 4 months in long COVIDs and asymptomatic matched controls compared to subjects infected with common cold coronavirus (HCoV) and unexposed healthy donors. B) Reduction of cytokine levels at 8 months in MC. IFN-β and IFN-λ1 level in LC were higher than MC at 8 months. Data shown as medians with interquartile ranges. Two-tailed *p* values <0.05 (*) were considered significant. Wilcoxon t-test was used for paired samples at 8 months.

**Figure 2.**
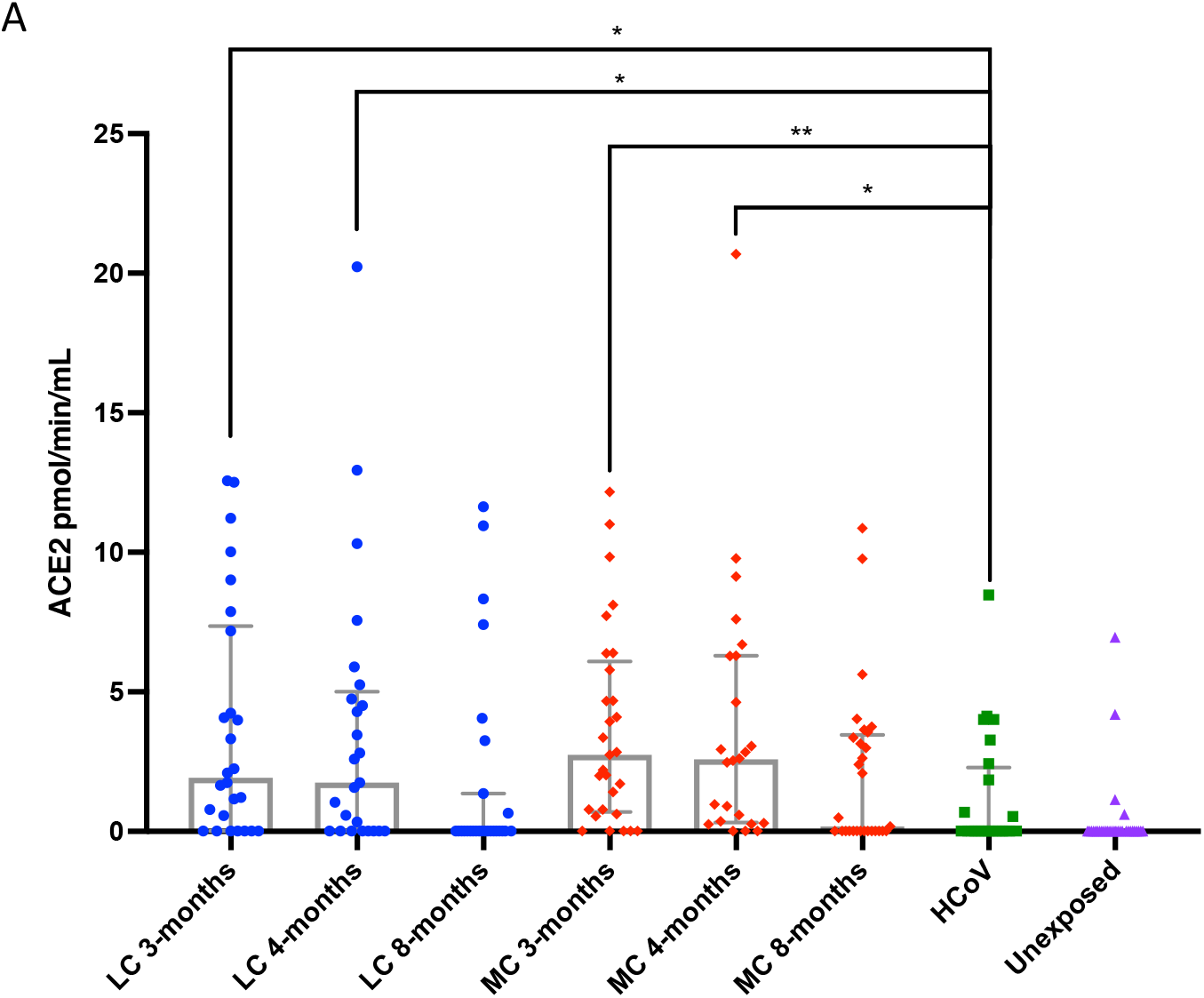
Elevated plasma ACE2 activity in SARS-CoV-2 infected subjects that decreases over time. A) Increased ACE2 activity at 3 months and 4 months post SARS-CoV-2 infection, that decreased at 8 months in both groups. Minimal ACE2 in HCoV and unexposed donors from University of Melbourne (Unexposed). LC (Long COVID), MC (asymptomatic matched controls). Data shown as column graphs showing medians with interquartile ranges. Two-tailed p values <0.05 (*) were considered significant.

To elucidate the persistence of these elevated analytes over a longer period of time, serum from all available subjects at 8 months were examined and compared with levels at 4 months (Fig. 1B). Decreases of both IFN-β and IFN-λ1 in MC group at 8 months were observed 4.4-fold (9.36[6.25-21.35] pg/mL, *p<0*.*0001*) and 1.8-fold (10.51[7.72-16.30] pg/mL, *p<0*.*05*) respectively. However this was not the case for LC group, where IFN-λ slightly decreased by 1.5-fold (22.32[18.75-49.07] pg/mL) and IFN-λ1 slightly increased by 1.05-fold (16.87[9.60-24.09] pg/mL) at 8 months, albeit not statistically significant. Overall, type I and type III interferons remained high in LC subjects and was significantly higher than the MC group at 8 months post-infection (*p<0*.*05*, for both interferons) and was higher than HCoV and unexposed groups (Suppl. Fig. 3A). Reduction of CXCL9 (MIG-1), CXCL10 (IP-10), IL-8 and sTIM-3 were evident in both groups at 8 months. There were also decreases in the levels of some of the 22 analytes not significantly different to control groups at month 4 by month 8 (Supp. Fig. 3B&C).

### Elevated levels of ACE2 in SARS-CoV-2 convalescent subjects

Plasma ACE2 activity levels have recently been shown to be elevated out to a median of 114 days following SARS-CoV-2 infection^25^, and we sought to investigate whether this was the case within the ADAPT longitudinal cohort at 3-, 4- and 8-months post-infection. At 3-months post-infection, median plasma ACE2 activity was significantly higher in both LC and MC groups compared to HCoV (LC=1.92 [0.02-7.36], MC=2.47 [0.63-5.94] *versus* HCoV =0.02 [0.02-1.99] pmol/min/mL, *p<0*.*01* and *p<0*.*001* respectively). These levels remained elevated at 4 months post-infection in both groups (LC=1.75 [0.02-5.01], MC=2.62 [0.36-6.3] *versus* HCoV =0.02 [0.02-1.99] pmol/min/mL, *p<0*.*05* and *p<0*.*001* respectively). Plasma ACE2 activity decreased at 8 months for LC (0.02[0.02-1.36] pmol/min/mL) and MC (0.18[0.02-3.46] pmol/min/mL). No significant difference was observed between these groups at 3, 4 and 8-months or when compared to HCoV. It appears that increased plasma ACE2 activity is specific to SARS-CoV-2 infection and is not a common feature of other coronaviruses.

### High inter-analyte correlations in long COVIDs at 4 months

Immune responses to viral infections involve communication of multiple specialised cell populations through direct interactions, secreted cytokines and various mediators in a concerted effort to combat the infectious agent. To obtain a broader scope of systems-level interactions during convalescence, we generated correlation matrices from 29 analytes measured at 4 months post-infection (Fig. 3). This analysis revealed multiple positive correlations across a spectrum of 18 markers including: IFN-α2, IFN-λ1, IFN-λ2/3 (interferons), IL-1β, IL-33 (IL-1 family cytokines); IL-12p70, MCP-1 (CCL2), IL-6, TGF-β1, CXCL-9 (myeloid activation), PD-1, sCD25 (T cell activation,) IFN-γ, TNF-α (Type 1 (Th1) cytokines); IL-5, IL-9, IL-13, (Type 2 (Th2) cytokines) and GM-CS in the LC group most of which were not present in the asymptomatic MC group. (Fig. 3 A&B). However at 8 months this profile had altered in both groups (Supp. Fig. 4A&B). This profile was not observed in HCoV (Supp. Fig. 4C) or unexposed donors (Supp. Fig. 4D).

**Figure 3.**
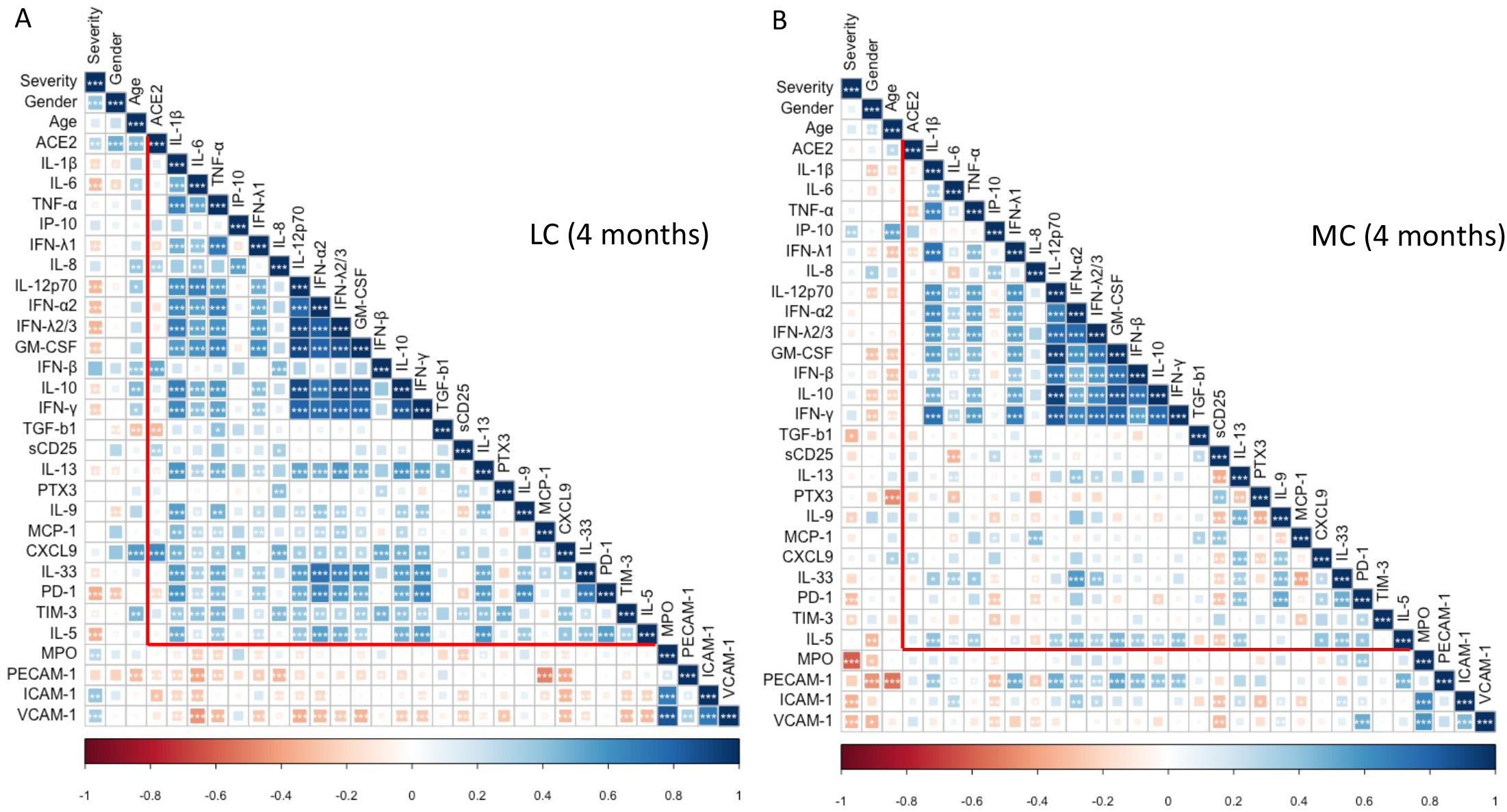
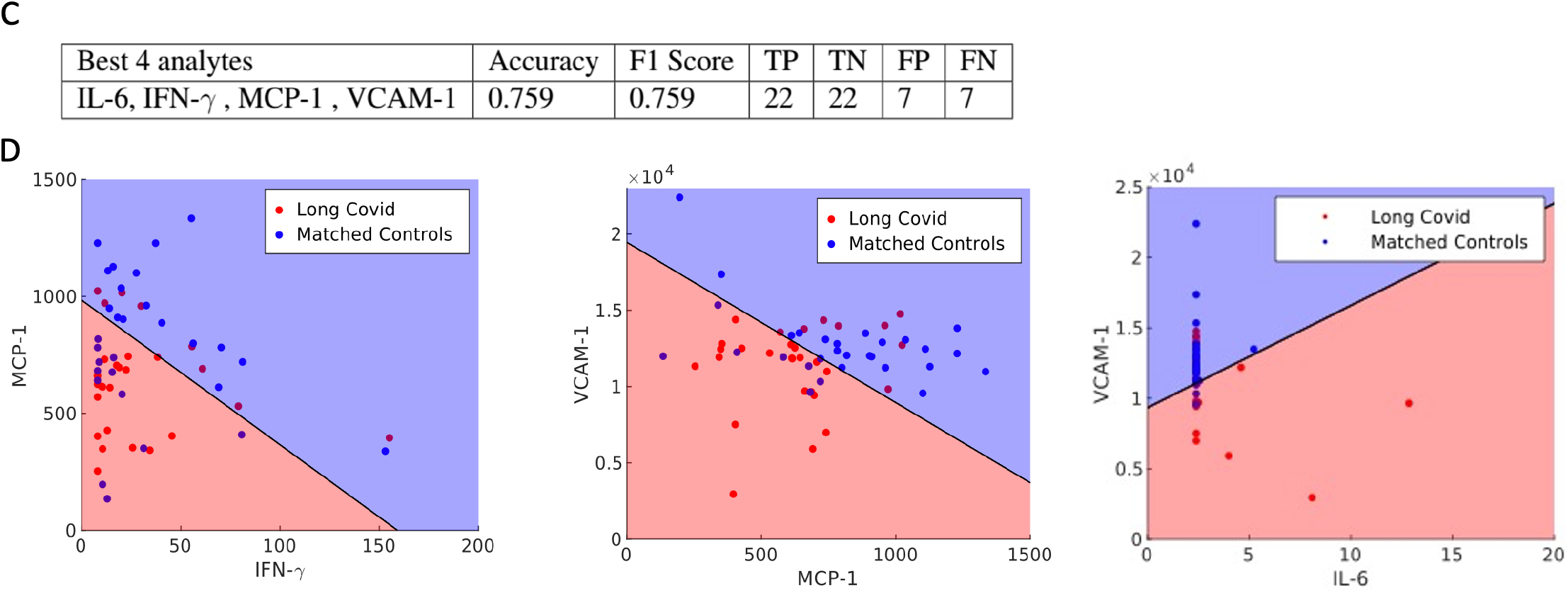
high inter-analyte correlations and the 4 best features in long COVID. A) Spearman correlation and hierarchical clustering comparing Severity, Gender, Ethnicity and Age with ACE2 and 28 other analytes at 4 months. Diffuse elevation of inflammatory analytes in Long COVIDs (highlighted in red triangle). B) No diffuse inflammatory profile in matched controls at 4 months. *p* values <0.05 were considered significant (*<0.05, **<0.01, ***<0.001). C) Best 4 analytes highly associated with LC eere: IL-6, IFN-γ, MCP-1 (CCL2) and VCAM-1. These four analytes gave an accuracy of 75.9%, and an F1 score of 0.759. TP= true positive, TN= true negative, FP= false positive and FN= false negative D) Visualisation of the domain boundary with two-dimensional projections. Red dots are long COVIDs, Blue dots are matched controls. Axes represent concentrations pg/mL.

**Figure 4.**
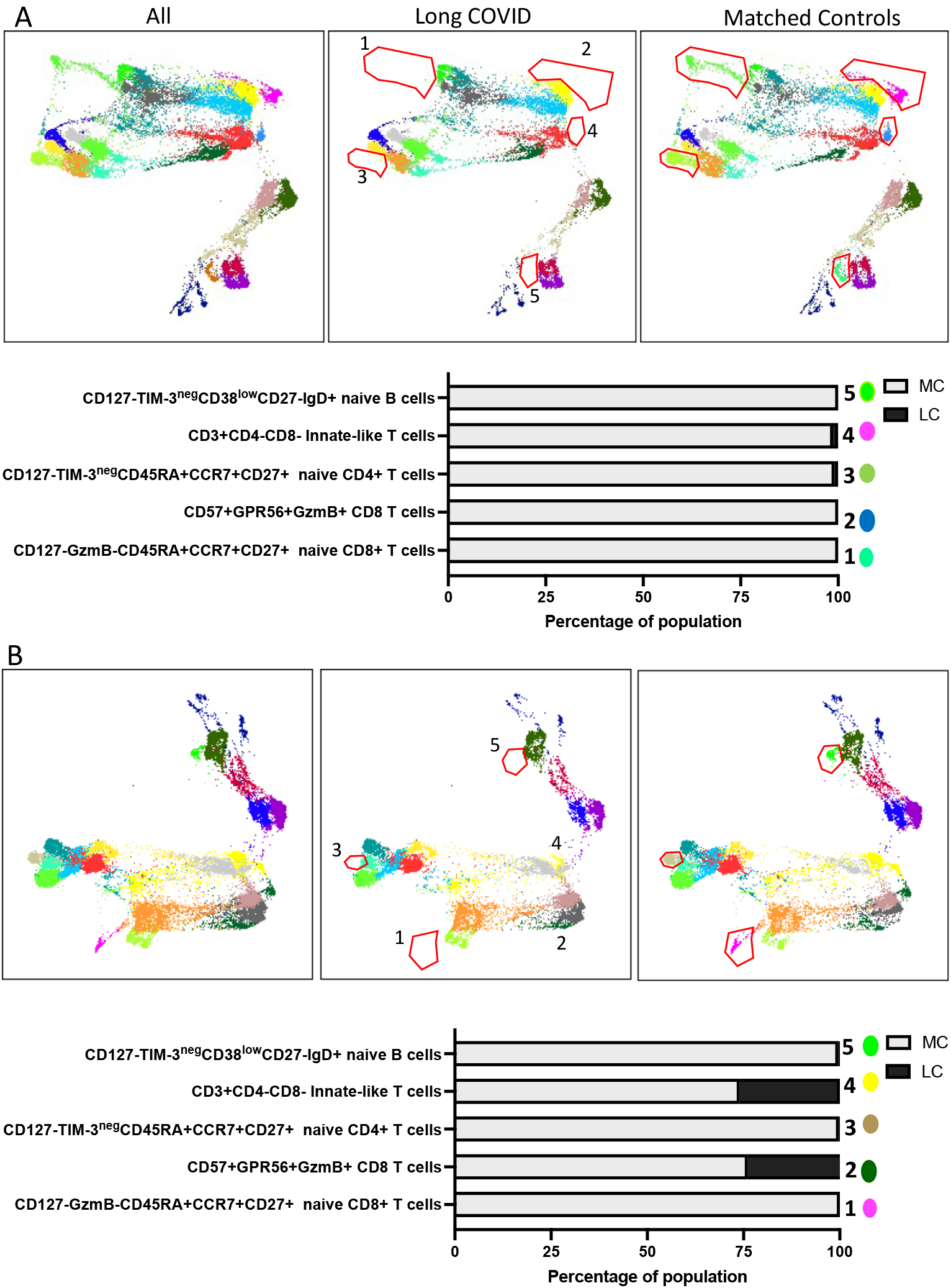
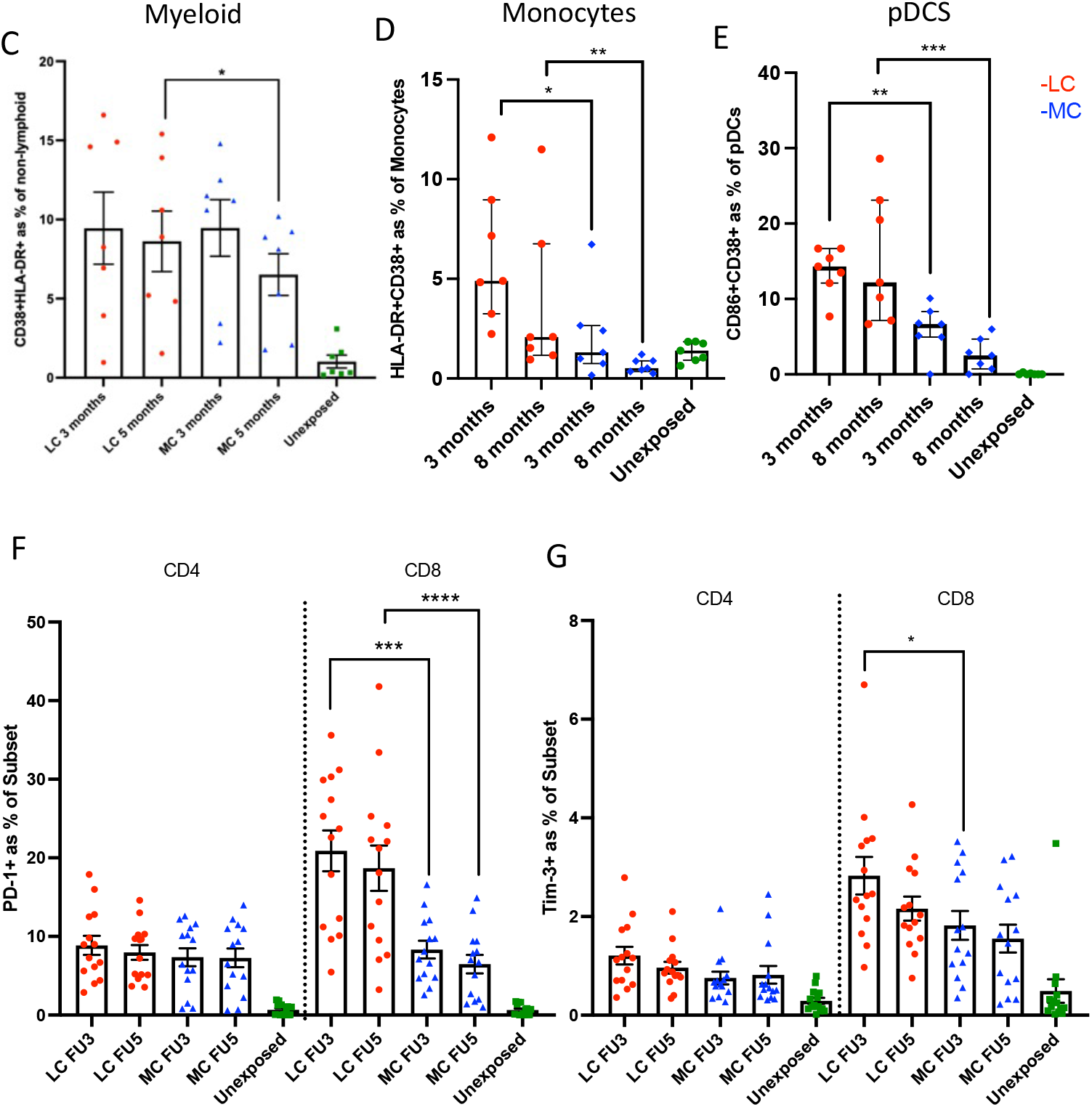
Distinct activation phenotype in non-lymphoid cells and absence of naïve T and B cells found in long COVID. A) Dimensional reduction utilising TriMap and clustering with Phenograph was used to visualise immune cell phenotypes. 5 populations consisting of un-activated naïve and cytotoxic phenotypes were absent in LC compared to MC at 3 months. Absent populations are outlined in red (middle panel), with frequency of population shown in bar graph. B) 3 naive populations remained absent in peripheral blood at 8 months in Long COVIDs. C) Activated non-lymphoid (myeloid) cells with combined expression of CD38 and HLA-DR in LC and MC at 3 months but was not reduce at 8 months in LCs. D) More activated monocytes and (E) plasmacytoid dendritic cells (pDCS) in LC compared to MC at 3 and 8 months. F) No difference in PD-1 levels on CD4 T cells, but higher expression by CD8+ T cells in LC at both timepoints. Data shown as medians with interquartile ranges. G) Higher Tim-3 expressed onCD8+ T cells in LC at 3months. Two-tailed *p* values <0.05 (*) were considered significant.

### Linear modelling defines 4 features most highly associated with long COVID

To identify a subset of the 29 analytes that are most strongly associated with long COVID we use linear classification. We trained our linear classifiers with data from 58 participants (29 with long COVID and 29 matched controls) with analyte concentrations after 4 and 8 months. The optimal number of analytes giving the highest accuracy when using a linear classifier to predict whether a participant suffered from long or asymptomatic COVID was 4. By exhaustively searching through all 20475 combinations of 4 analytes we found that the 4 analytes IL-6, IFN-γ, MCP-1 (CCL2) and VCAM-1 gave an accuracy of 75.9%, and an F1 score of 0.76 (Fig. 3C). Beyond identifying the four analytes that are most highly associated with long COVID, linear classifiers offer a simple test to predict whether or not a patient will suffer from long COVID. Linear classifiers define what is known as a decision boundary. A patient’s 4 analyte concentrations at 4 and 8 months will lie on either side of this boundary, and which side it is will determine whether or not the patient is predicted to experience long or asymptomatic COVID. The decision boundary for the 4 best analytes is eight-dimensional due to both the 4- and 8-month data. However, we can visualise the domain boundary with two-dimensional projections of IFN-γ, against MCP-1, MCP-1 against VCAM-1 and VCAM-1 against IL-6 as shown in Fig, 3D.

### Highly activated innate cells combined with absence of un-activated naïve T and B cell subsets-a distinguishing feature of long COVID

To investigate differences in immune cell phenotypes between LC and MC, a 19-parameter phenotyping panel was developed and utilised for PBMC samples from LC and MC groups at 3 months and 8 months following infection, with MC group being the primary comparator. Dimensional reduction via TriMap coupled with Phenograph clustering identified 24 distinct cell clusters at month 3 and 21 clusters at month 8 (example of cell cluster identification shown in Suppl. Fig. 5A); comprising T cells, B cells, NK cells and non-lymphoid (myeloid) cells (Supp. Fig. 5B&C). Of the 24 subsets identified at month 3, five were absent in LC subjects (Fig. 4A), these include:-1) CD127^low^GzmB-CCR7+CD45RA+CD27+ naïve CD8+ T cells, 2) CD57+ highly cytotoxic (GPR56+GzmB+) CD8+ T cells, 3) CD127^low^TIM-3^neg^CCR7+CD45RA+CD27+ naïve CD4+ T cells, 4) CD3+CD4-CD8-innate like T cells (may comprise NKT cells and γ δ -T cells), and 5) naïve CD38^low^CD27-IgD+ B cells. At 8 months only 3 populations remined absent: 1) CD127^low^GzmB-CCR7+CD45RA+CD27+ Naïve CD8+ T cells, 2) CD127^low^TIM-3^neg^CCR7+CD45RA+CD27+ Naïve CD4+ T cells, and 3) CD38^low^CD27-Naïve IgD+ B cells (Fig. 4B). These findings are indicative of ongoing inflammation through to 8 months in LC subjects. There was no reconstitution of these un-activated naïve T and B cell subsets even at 8 months post-infection in peripheral blood, however other naïve subsets that appear more activated are present in LC. All concatenated patients sampled (7 LC, 7 MC and 7 unexposed) contributed to all populations identified (Suppl. Fig 6A-D). These un-activated T and B cells are evident in unexposed and MC comparator groups.

Highly activated non-lymphoid cells (defined by the co-expression of CD38+HLA-DR+) remained elevated at 8 months in LC (Fig. 4C). In contrast, activation markers on non-lymphoid cells decreased over time in asymptomatic matched controls (median 3 months= 11.2[3.43-12.5] and 8 months= 8.12[2.07-9.23], *p<0*.*05*). Activate monocytes (CD14+CD16+) were higher in LC compared to MC at both 4 and 8 months (3.74-fold, *p<0*.*05*, and 4-fold, *p<0*.*01*, respectively) (Fig. 4D). Plasmacytoid dendritic cells expressing activated markers CD86+ CD38+ were also higher in LC at both timepoints (2.14-fold, *p<0*.*01* and 4.88-fold, *p<0*.*001*, respectively) (Fig. 4E). No differences in activation of myeloid dendritic cells were observed (Suppl. Fig. 7A). Notably, the immune checkpoint receptor commonly associated with T cell activation/exhaustion PD-1 was highly expressed on CD8+ T cells at 3 and 8 months in LC compared to MC (3.04-fold, *p<0*.*001* and 2.86-fold, *p<0*.*0001*)(Fig. 4F). T cell exhaustion marker Tim-3 was also higher in LC at month 3 (1.6-fold, *p<0*.*05*), but decreased at month 8 (Fig. 4G). However, no difference was observed between LC and MC as judged by co-expression of PD-1 and Tim-3 on CD4+ and CD8+ T cells (suppl. Fig. 7B).

## Discussion

Our study demonstrates several unique and important findings in relation to understanding long-term outcomes from COVID-19. We show that convalescent profiles post COVID-19 are different from other coronaviruses. Several cytokines, mostly type I and III interferons, but also chemokines down stream of IFN-γ, were highly elevated in people following resolution of active SARS-CoV-2 infection compared to HCoV and unexposed controls at 4 months. IFN-γ and IFN-λ1 remained high in long COVID to 8 months after initial infection, while the levels of these two cytokines had begun to resolve in the matched controls. This finding was observed irrespective of persistent long COVID symptoms. We also demonstrated an elevation of plasma ACE2 levels at 4 months post COVID infection with the degree of elevation not being different between those with and without long COVID symptoms. ACE-2 levels trended towards normal by 8 months. Further, by employing a linear classification model we identified 4 biomarkers highly associated with long COVID: IL-6, IFN-γ, MCP-1 (CCL2) and VCAM-1. This indicates that components of the acute inflammatory response, T cell, Myeloid cell and vascular endothelium activation are associated with long COVID. Interestingly, immune cell phenotyping revealed chronic activation of: a subset of CD8 T cells with expansion of PD-1 and Tim-3 positive subsets; plasmacytoid dendritic cells; and monocytes (CD38 and HLA-DR co-expression) in long COVID patients. Levels of these subsets are increased in long COVID patients relative to matched controls at 3 months with perturbations persisting out to 8 months. Further, there is an apparent turnover of un-activated naïve T and B cell subsets in peripheral blood in long COVID subjects over the same time period. This suggests maintenance of long-lasting inflammation and immune activation in these individuals. Together these findings suggest that SARS-CoV-2 infection exerts a unique residual prolonged effect on aspects of both the innate and adaptive immune systems, and that this may be driving the post COVID symptomology known as ‘long COVID’.

Type I and III Interferons, namely IFN-β and IFN-λ1 were highly elevated in our COVID-19 convalescent subjects compared to HCOV and unexposed subjects,. While their levels decreased over time in recovered subjects they remained high in those with long COVID. IFN-β can be expressed by most cell types and can be induced early during viral infection, before most IFN-α subtypes ^26-28^. IFN-λ1 or IL-29 can also be induced in a broad range of cell types including epithelial cells by stimulation of toll-like receptors (TLRs), Ku70, and RIG-1-like receptor ^29^, however, as for type I interferons, the main producers were found to be type 2 myeloid dendritic cells (plasmacytoid DCs)^30-32^. Type I interferon receptors are widely expressed on immune cells, and IFN-β responses during influenza and SARS-CoV-1 infection can result in immunopathology ^33,34^. In contrast, IFN-λ receptors are mainly expressed at epithelial barriers and confer localised antiviral protection ^35,36^. The morbidity of acute COVID-19 infection appears to correlate with high expression of type I and III interferons in the lung of patients ^37^. Furthermore IFN-λ produced by dendritic cells in lungs of mice in response to synthetic viral RNA is associated with damage to lung epithelium ^38^, and IFN-λ signalling can hamper lung repair during influenza infection of mice ^39^. Diminished type I interferon and enhanced IL-6 and TNF-α responses in severe acute COVID-19 patients were reported by Hadjadj *et al* ^38^. Our cohort of long COVID subjects, however, mostly consisted of patients with mild and moderate initial illness, and maintenance of elevated type I and III interferon levels at 8 months demonstrates ongoing inflammation within these individuals and is consistent with our observation of prolonged activation of plasmacytoid dendritic cells.

CXCL9 (MIG-1) and CXCL10 (IP-10) are pro-inflammatory T cell chemo-attractants that are expressed by monocytes, endothelial cells, and fibroblasts downstream of interferon-gamma signalling ^40^. Upon engagement with their receptor CXCR3, mainly expressed on T cells, they drive naïve CD4+ T cell differentiation into effector Th1 cells, as well as recruiting CD4+ and CD8+ T cells in sites of tissue damage, which has been documented during vascular disease and pulmonary fibrosis ^41,42^. Both chemokines have been implicated in symptomatic acute coronavirus infections, including COVID-19, where levels were higher in severe compared to asymptomatic subjects ^43^. Similarly, CXCL8 (IL-8) produced by lung epithelium and airway resident macrophages, has previously been shown to be released by lung cells after activation by spike proteins from SARS-CoV-1 ^44^. CXCL8 acts as a chemoattractant that facilitates migration of inflammatory T cells and neutrophils, that may contribute to alveolar damage ^45,46^. Further the elevation of CXCL9 (MIG-1) and CXCL10 (IP-10) is consistent with persistent absence of un-activated naïve cell subsets in peripheral blood that we observed in the long COVID group.

Increased shedding or cleavage of ectodomains of cell surface proteins during inflammation has been well documented ^47,48^. Increased levels of soluble T-cell immunoglobulin mucin domain-3 (sTIM-3) a marker of T-cell activation and exhaustion has been reported in chronic viral infections (i.e., HIV, hepatitis B and C)^49-51^ where there is persistent antigenic stimulation, and recently in COVID-19 infection, where patients admitted to ICU had high plasma levels of sTIM-3 and myeloperoxidase (MPO) ^52^. Elevated sTIM-3 levels were seen in those with long COVID, but not recovered subjects or those with other HCoV, and is entirely consistent with our observation of expanded subsets of memory CD8 T cells expressing Tim-3 and PD-1 in the same group who appear to have chronic T cell activation and potentially T cell exhaustion perhaps partially driven by the prolonged presence of activated antigen presenting cells and the downstream drivers of T cell activation such as type I interferons and the IFN-γ driven chemokines CXCL9 and CXCL10. Another membrane bound protein found to be ‘shed’ during COVID-19 is ACE2, which is also an important receptor for viral host cell entry ^53^. Patel *et al* showed that shedding of ACE2 from the cell membrane resulted in increased plasma ACE2 activity levels in convalescent COVID-19 subjects ^25^. Our data corroborates with this finding, where elevated plasma ACE2 activity occurred in COVID-19 infection regardless of symptom severity, at a higher level than HCoV and unexposed, persisted at 4 months, and decreased at 8 months in most subjects.

We observed high inter-analyte correlations in long COVIDs at 4 months that was not observed in MC, HCoV or unexposed comparator groups. These correlations reinforced the idea that there was interrelated activation of acute phase inflammatory, innate and adaptive immune responses. This profile was less prominent at 8 months. In order to assess across all combinations of analytes those most strongly associated with long COVID, a linear classification model was employed. A combination of 4 biomarkers, IL-6, IFNγ, MCP-1 (CCL2), and VCAM-1 was identified as being the most accurate correlate of long COVID. These analytes could be more prominent at the acute stage of infection. IL-6 is a pleiotropic mediator that has an effect on inflammation and immune activation ^54^. IFNγ produce by NK and T cells have anti-viral activity and levels are associate with a heightened inflammatory state. High IL-6/IFN-gamma ratio was found to be associated with severe disease in COVID-19 patients at acute infection ^55^. MCP-1 (CCL2) is a chemokine that is produced by many cell types (endothelial, fibroblasts, epithelial, monocytic etc ^56^) and acts as a chemoattractant for monocyte, NK, neutrophils, basophils, B and T cells and is involved in T cell activation ^57^. VCAM-1 mediates adhesion of immune cells to the endothelium and has been implicated in inflammatory processes ranging from atherosclerosis and rheumatoid arthritis ^58^. Recent meta-analysis of > 300,000 single-cell transcriptomic profiles from COVID-19-affected lungs have found that IFNγ together with TNFα drive an drive a IP-10+CCL2+ macrophage phenotype expanded in severe COVID-19 lungs and inflammatory diseases with tissue inflammation ^59^. These analytes could be more prominent at the acute stage of infection and warrants further investigation. The observation that the best correlate of the presence of long COVID is an eclectic combination of biomarkers reinforces breadth of host response pathways that are activated during long COVID.

Finally, T cell activation via surrogate markers CD38 and HLA-DR, T cell exhaustion, together with an increase in B cell plasmablasts during severe COVID-19 has been reported ^60-62^. We found no difference between activation levels as judged by cell surface expression of these molecules on T cells between long COVID and asymptomatic (data not shown) but identified highly activated myeloid cells comprising monocytes and dendritic cells. Elevated levels of CD38+HLA-DR+ myeloid cells decreased over time within asymptomatic subjects but remained unchanged in those with long COVID. It is known that type I and type III interferons upregulate MHC-class expression including HLA-DR ^63^. When an unbiased large-scale dimensional reduction using triplets (TriMap) approach was used, we identified 5 population clusters that were absent in those with long COVID. These consisted of mostly un-activated naïve T and B cell subsets (CD4 (CD127^low^TIM-3^neg^CCR7+CD45RA+CD27+), CD8 (CD127^low^GzmB-CCR7+CD45RA+CD27+) and CD127^low^TIM-3^neg^CD38^low^CD27-IgD+ B cells), as well as double negative (CD4-CD8-) Innate-like T cells and highly cytotoxic CD57+GPR56+GzmB+ CD8+ T cells at 3 months. The depletion of these un-activated naïve subsets persisted at the 8-month time-point. Other more activated naïve subsets were present at 3 months in both groups and remained at 8 months. Naïve T and B cell expressed low to mid-levels of CD127 in MC and unexposed (Suppl. Fig. 6E&F), but CD127^low^Tim-3 ^neg^ phenotype was absent in LC. Taken together these observations suggest persistent recruitment of activated naïve T cells, potentially due to by-stander activation secondary to underlying inflammation and/or antigen presentation by activated plasmacytoid dendritic cells or monocytes, pushing naïve cells into an activated state, thereby depleting the un-activated naïve T and B cell pools in those with long COVID. The ultimate result of this chronic stimulation may be the observed expansion of exhausted PD-1 or Tim-3 positive CD8+ T cells. Bystander activation of un-activated naïve subsets into more activated phenotypes is consistent with observations in subsets of patients during acute severe COVID-19 infection ^64,65^.

Our data does have some limitations. Firstly, although our long COVID ‘cases’ and controls were matched for two major characteristics (age and gender) it is possible that the differences observed reflect other key demographic or other factors between the groups. While long COVID group had a higher representation of those with severe acute disease (8 LC and 2 MC), sensitivity analyses excluding these patients did not substantially alter the statistical significance any of our major associations. Also, due to timing of ethics approval and trial setup we were not able to collect samples at acute infection. We are therefore unable to differentiate whether the elevations in biomarkers seen during convalescence correlate with levels during the acute infection. A review of the literature of biomarkers during acute infection suggests that while some of the alterations observed here are potentially consistent with a hypothesis that the major drivers of the levels of biomarkers in convalescence are those in the acute infection, others are not. For instance diminished type I interferon and enhanced IL-6 and TNF-α responses in severe acute COVID-19 patients have been reported by Hadjadj *et al* ^38^. Our cohort consisted mostly of patients with mild and moderate initial illness and among our cohort those with long COVID do not have elevated levels of IL6 or TNF, but do have elevated levels of both type I and III interferon. Although 7 of the 29 serum analytes were abnormally elevated, key cytokines implicated in other fatigue syndromes, such as Pentraxin-3(PTX3), TGF-β1, and IL-13^66,67^, were not. This suggests long COVID may be differentiated from other post viral fatigue syndromes, however, our results require validation in other cohorts of long COVID to ensure their replicability. Finally, our definition of ‘long COVID’ cases was internally set given the lack of international consensus regarding this and thus is not definitive. Nevertheless, the inclusion of three of the commonest persisting symptoms and the blinding of cases and controls from the laboratory scientists helps to ensure the validity of our findings. Additional strengths of our study include the prospective protocol defined collection of samples and data and the establishment of an ADAPT–C cohort of individuals affected contemporaneously with other human coronaviruses to act as comparators.

In summary, our data strongly suggest there is an ongoing, sustained inflammatory response following even apparently mild to moderate acute COVID-19 infection, which is not found following infections with other prevalent coronaviruses. This process may be asymptomatic and therefore subclinical and takes more than 8 months to resolve. In those with persistent long COVID symptoms as distinct from asymptomatic convalescence, there is a sustained increase, out to at least 8 months, in highly activated myeloid cells, particularly monocytes and plasmacytoid dendritic cells, that likely contribute to the sustained elevation of inflammatory cytokines (particularly type I and III interferons), despite the majority of cohort suffering initial mild-moderate illness. Further, there was evidence of sustained activation of the adaptive immune system manifesting in perturbations of both naïve and memory/effector cell compartments with absence of the un-activated naïve T and B cells and increases in the expression of checkpoint molecules PD-1 and Tim-3 on CD8+ memory T cells. This widespread activation signature is also reflected in the identification of serum levels of IL-6, IFN-γ, MCP-1 (CCL2), and VCAM-1 as the 4 features that most strongly associated with long COVID. The drivers of this activation require further investigation but possibilities include persistence of antigen, an autoimmune phenomenon driven by unexpected antigenic cross reactivity, or a reflection of damage repair. To our knowledge, this is the first demonstration of an abnormal immune profile in COVID-19 patients at extended time points post infection and provides clear support for the existence of a syndrome of ‘long COVID’. While our findings require validation in other similar cohorts, they provide an important foundation to understanding further the pathophysiology of this syndrome and potential therapeutic avenues for intervention.

## Data Availability

Source data files will be uploaded upon submission to intended journal

## Acknowledgements

The authors thank the staff at the St Vincent’s Institute for Applied Medical Research-Clinical Trials Unit for their expertise in specimen processing and bio-banking. We appreciate grant support from the St Vincent’s Clinic Foundation, the Curran Foundation, the Rapid Response Research Fund (UNSW) and the Medical Research Futures Fund (Australia). SKP and LMB-NHMRC programme grant APP 1055214 (LMB), Medical Research Future Fund award GNT 1175865, Austin Medical Research Foundation Grant. DBW-NSF-DMA grant 1902854, SJK and JAJ-the Victorian Government, MRFF Award (2005544), NHMRC program grant (1149990 (SJK, ADK), NHMRC Fellowships (1136322-SJK, 1123673-JAJ).

---

**Supplementary Table 1.**
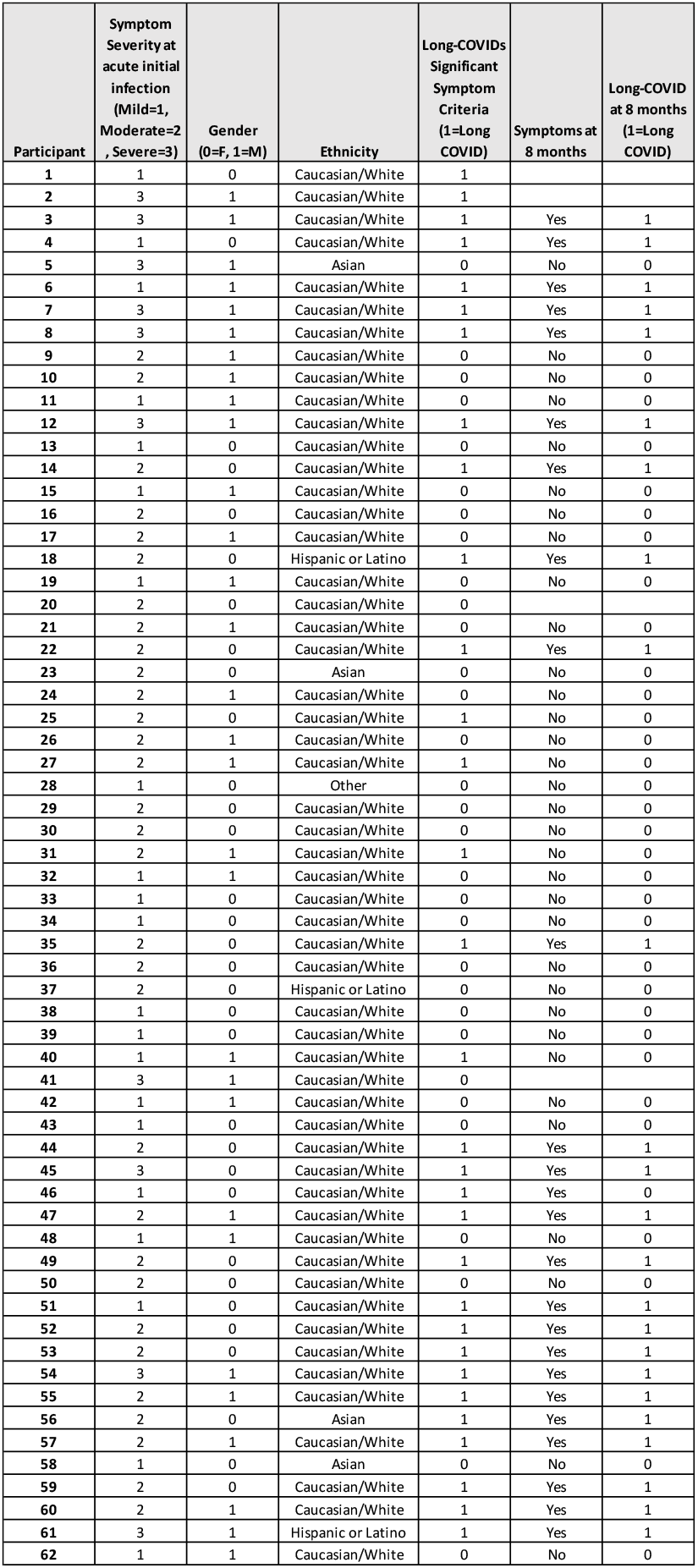
Clinical data of Long COVID and Matched controls. A) Symptom severity, Gender, Ethnicity of 31 long COVID and 31 asymptomatic matched controls. Subject 15 was on Everilomus and Mycophenolate due to prior renal transplant, and subject 45 was on Plaquenil for treatment of SLE.

**Supplementary Table 2.**
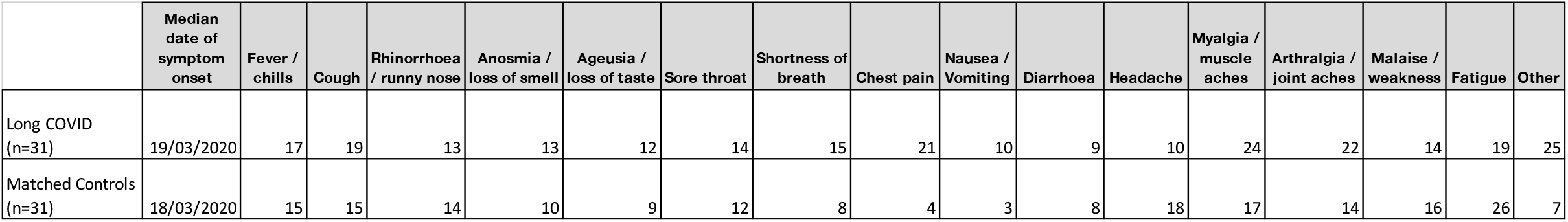
Symptoms at acute initial infection. A) Median date of symptom onset for two cohort groups (long COVID and matached control). Numbers represent subjects with reported symptom at acute initial infection.

**Supplementary Figure 1.**
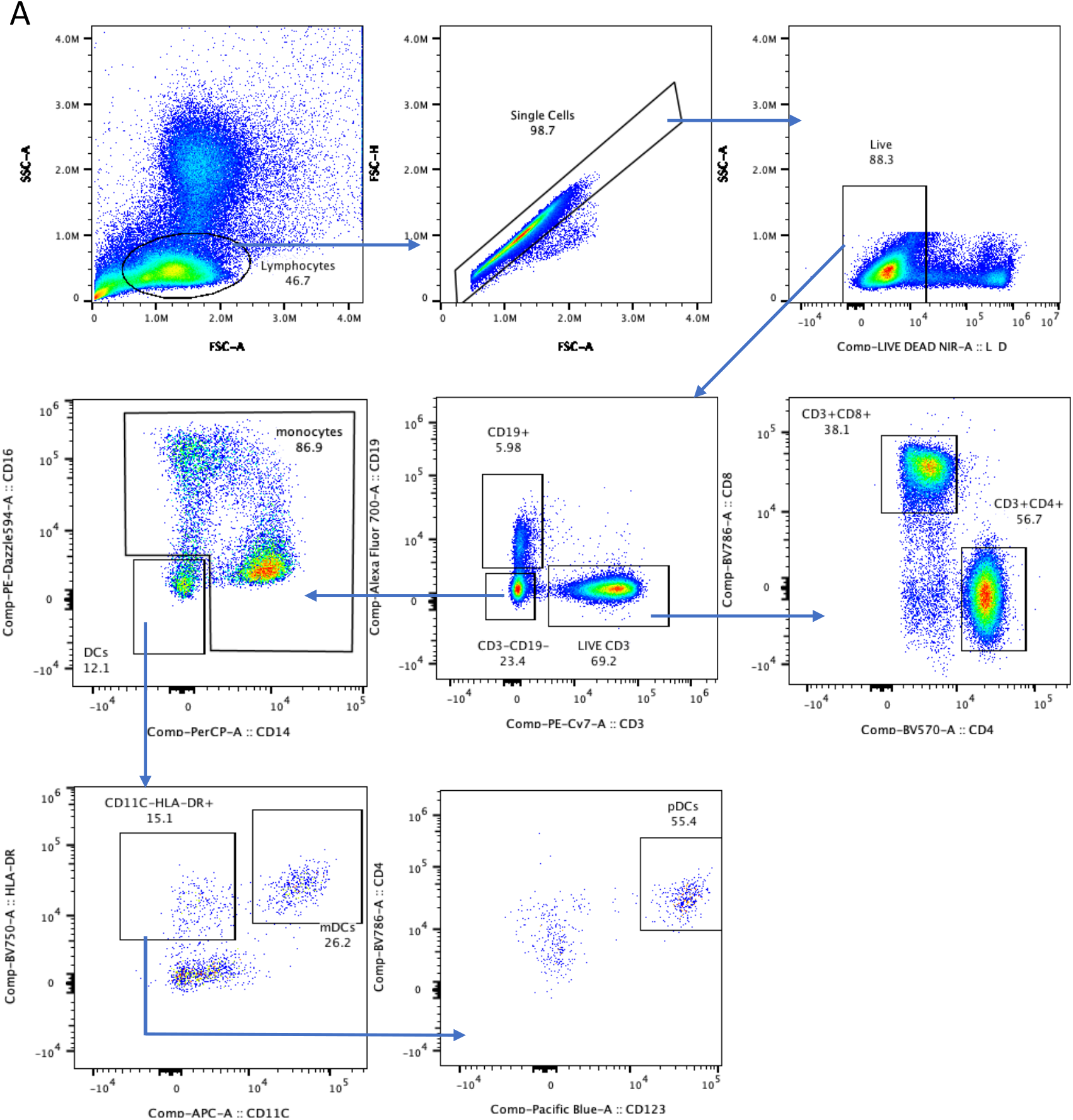
Gating strategy. A) Representative dot plot and gating strategy for flow cytometry analysis. Cells use for downstream analysis were live singlet expressing lineage markers, mDC (myeloid dendritic cells), pDC (plasmacytoid dendritic cells.

**Supplementary Figure 2.**
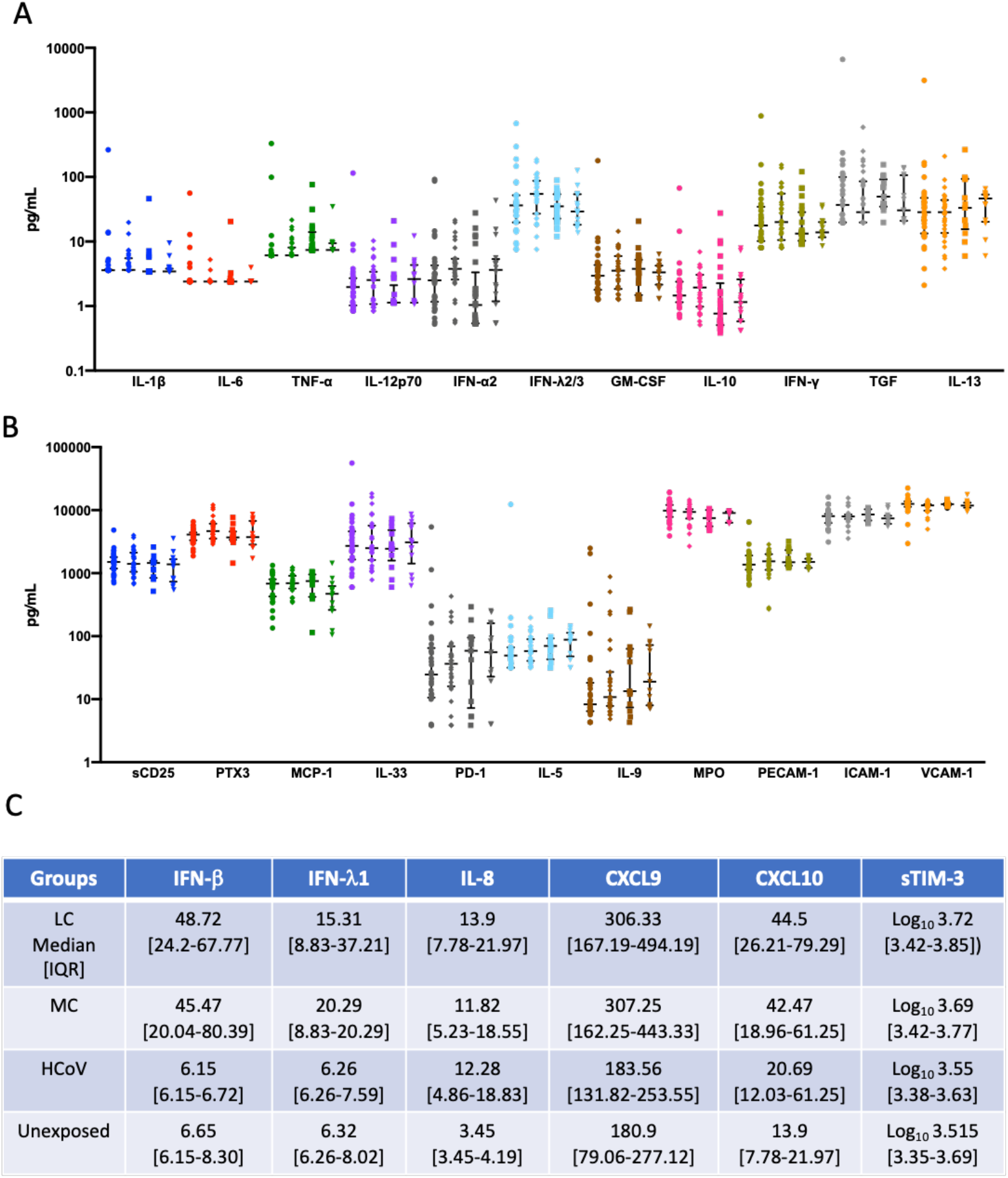
No difference in levels of 22 analytes following SARS-CoV-2 infection. A) No differences in IL-1b, IL-6, TNF-α, IL-12p70, IFN-α2, IFN-λ2/3, GM-CSF, IL-10, IFN-γ, TGF-β1 or IL-13 levels between the 4 groups at 4 months. B) No differences in sCD25, PTX3, CCL2(MCP-1), IL-33, PD-1, IL-5, IL-9, MPO, PECAM-1, ICAM-1 or VCAM-1 levels between the 4 groups. Data shown as medians with interquartile ranges. C) Median and IQR values for 4 groups at 4 months.

**Supplementary Figure 3.**
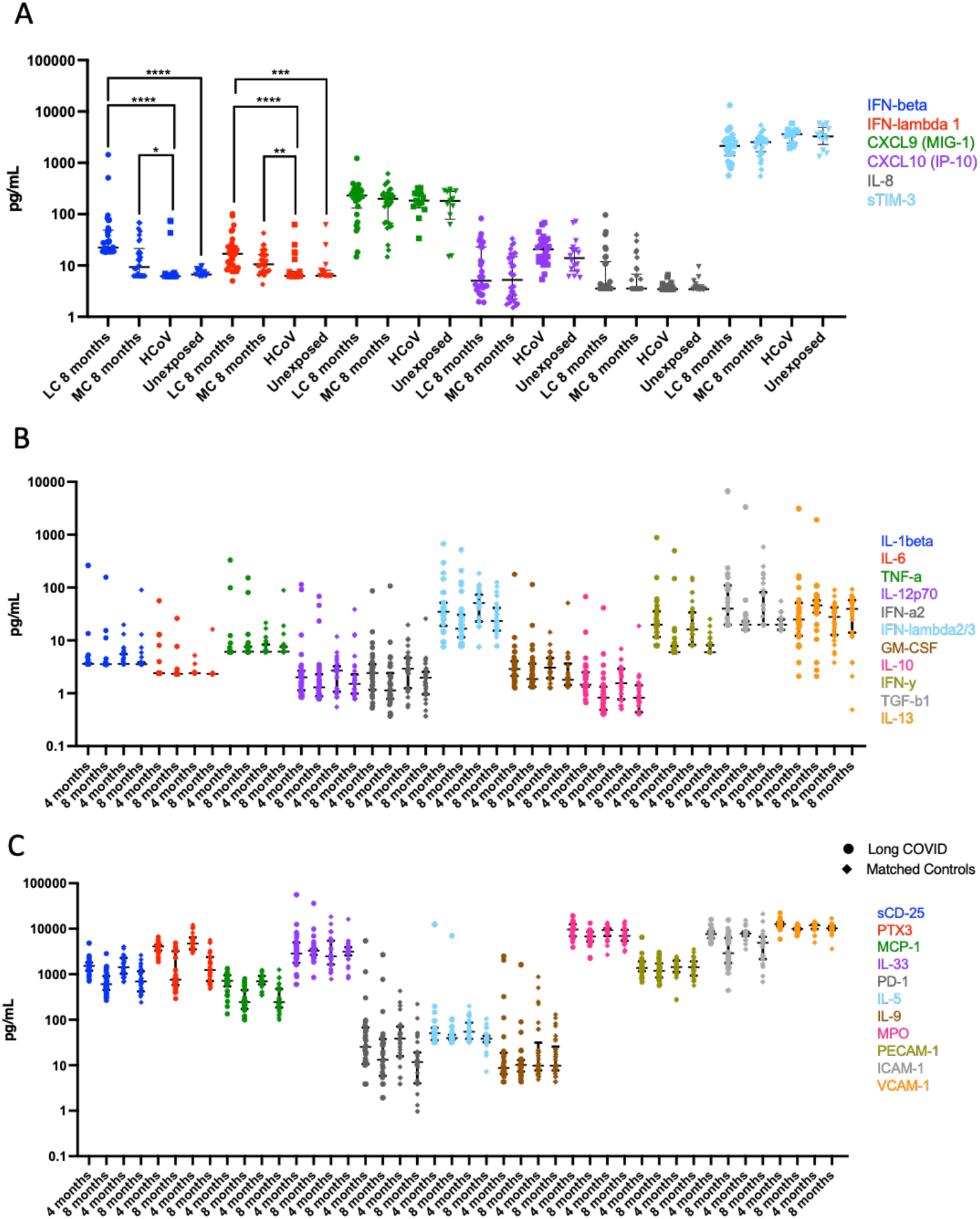
Reduction of some cytokines at 8 months. A) IFN-β and IFN-λ1 levels remained higher in LC at 8 months compared to HCoV and unexposed. B) Significant reduction of TGF-β1 at 8 months in LC. C) Decrease in most analytes at 8 months. Data shown as medians with interquartile ranges.

**Supplementary Figure 4.**
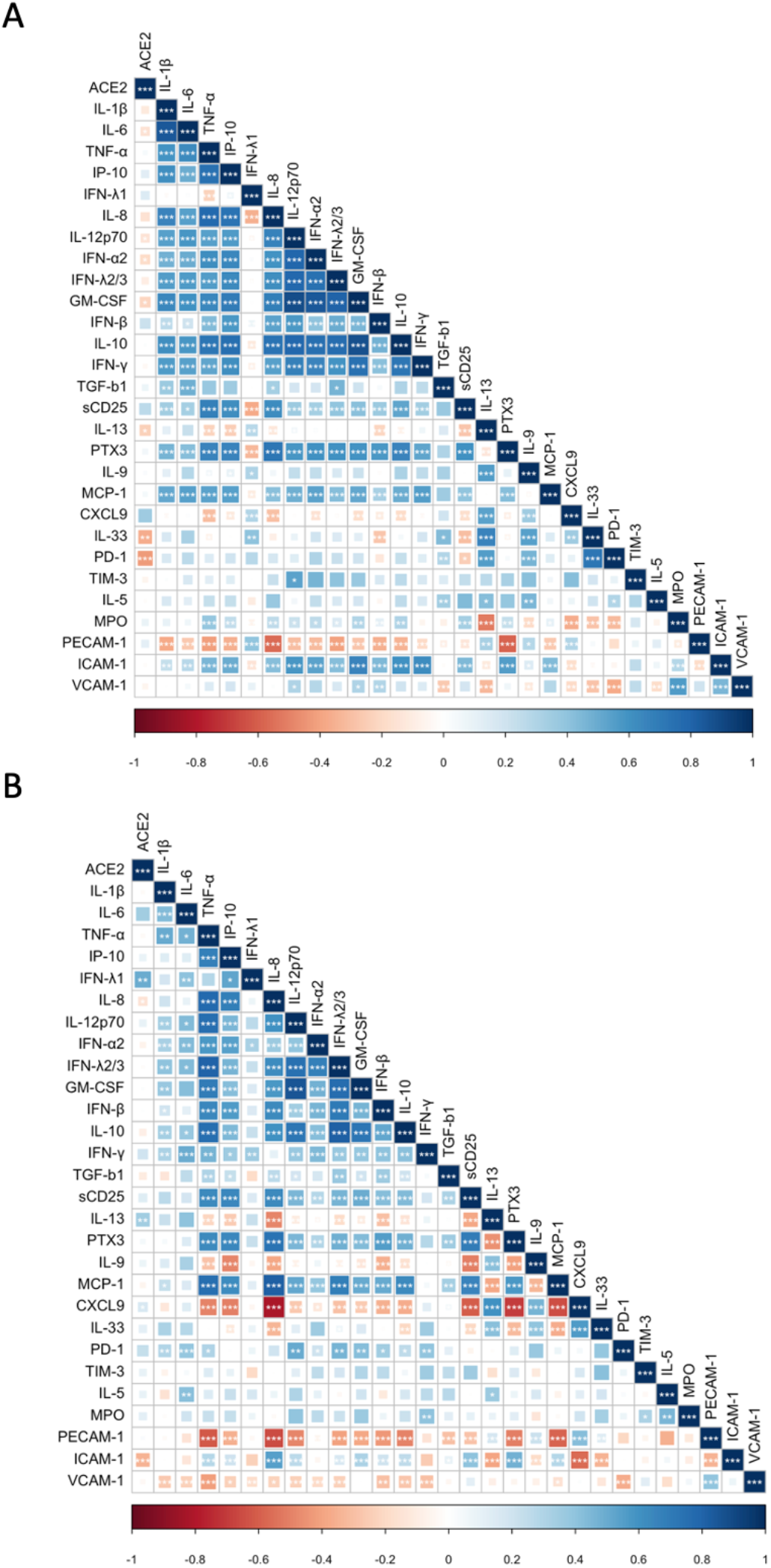
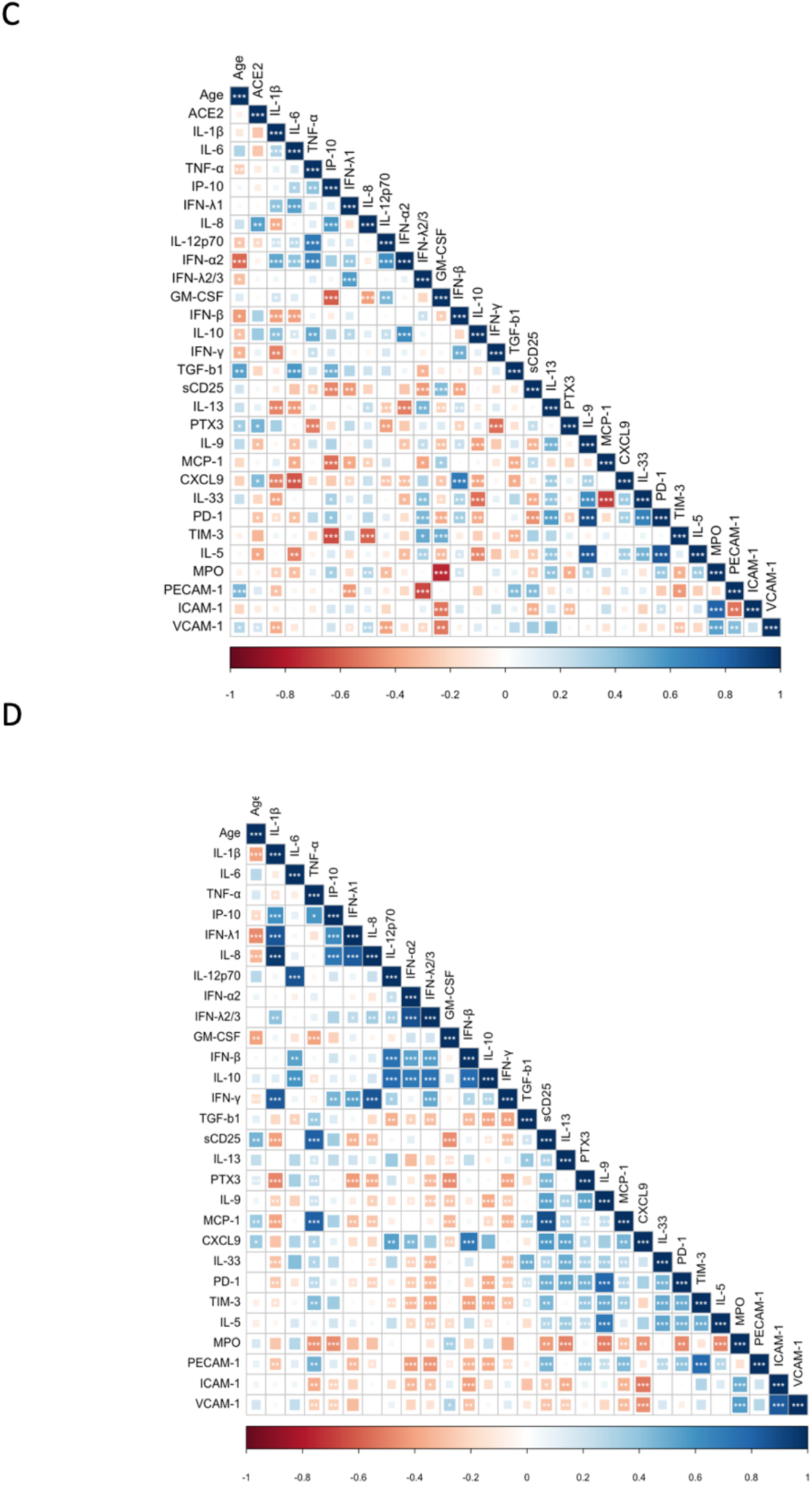
Correlation matrices from HCoV and unexposed donors. Spearman correlation comparing indicated features in LC (A) and MC (B) at 8months and common cold coronavirus infected donors-HCoV (C) and unexposed healthy donors (D).

**Supplementary Figure 5.**
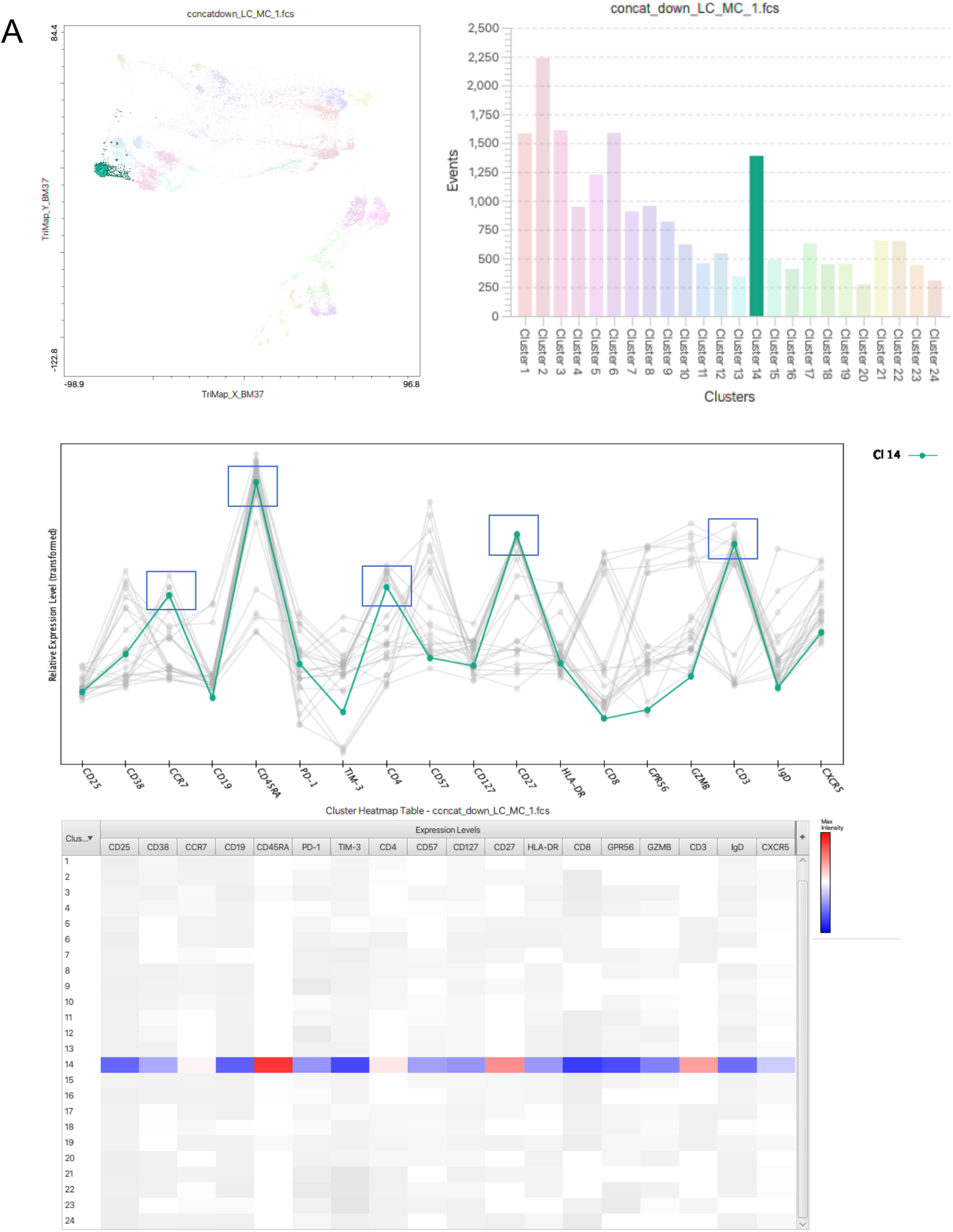
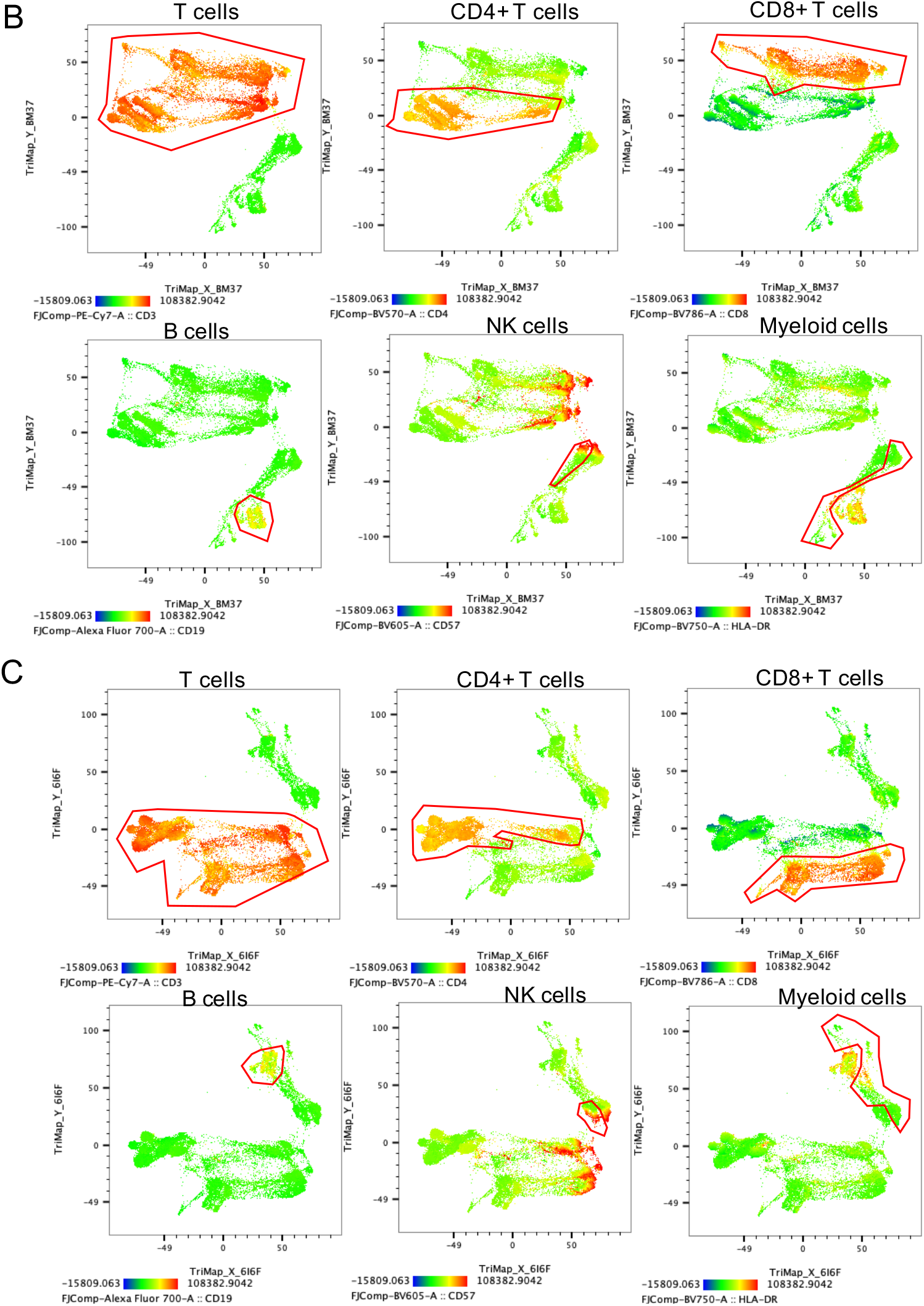
Clus tering of cell phenotypes. *A)* Example of cell cluster phenotype identification using CellExplorer plugin on Flowjo. Cluster 14 is equivalent to population 3, identified as CCR7+CD45RA+CD27+ naïve CD4+ T cells. TriMap of T cell, B cell, NK Cell and myeloid cell clusters (outlined in red) at 3 months (B) and 8 months (C).

**Supplementary Figure 6.**
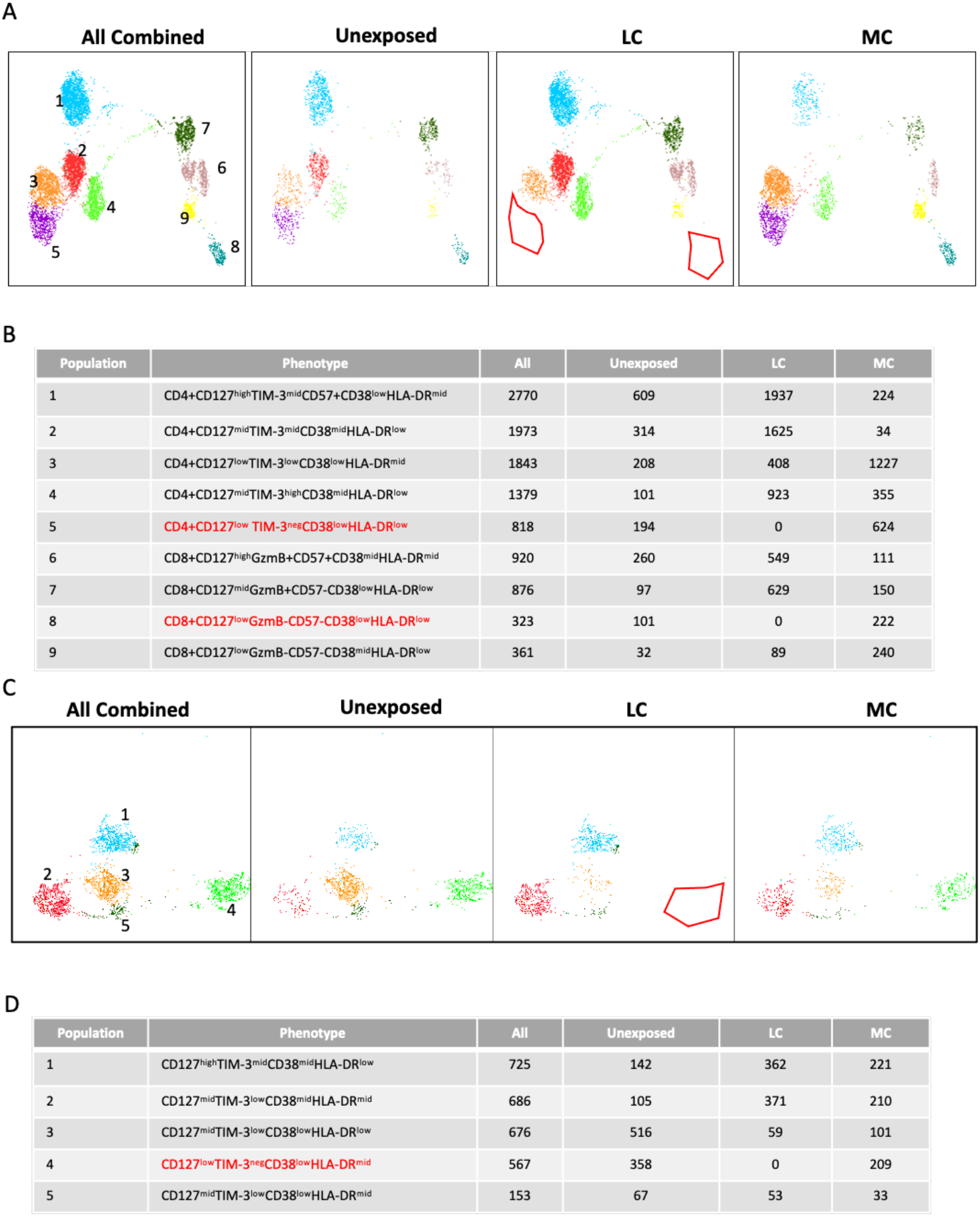
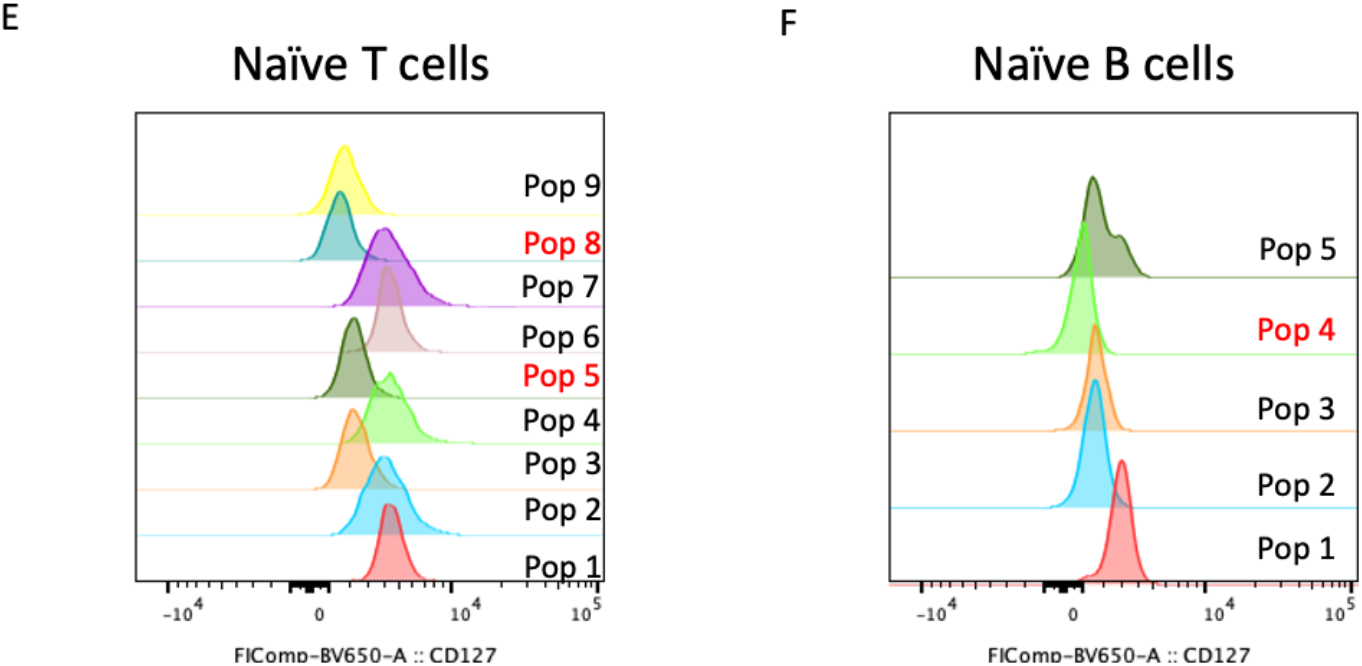
Deciphering naïve phenotypes. Representative TriMap and phenograph clustering of CCR7+CD45RA+ naïve T cells (A) and lgD+CD27- naïve B cells (C). Red gates and text represent un-activated naïve subsets absent in long COVID subjects. Phenotype and cell frequencies from naïve T cell s (B) and naïve B cells (D). CD127 expression on differing naïve T cell (E) and naïve B cell (F) populations (all data combined). Red text highlights subsets absent in long COVID. Unexposed, LC (long COVID0, and MC (asymptomatic matched controls).

**Supplementary Figure 7.**
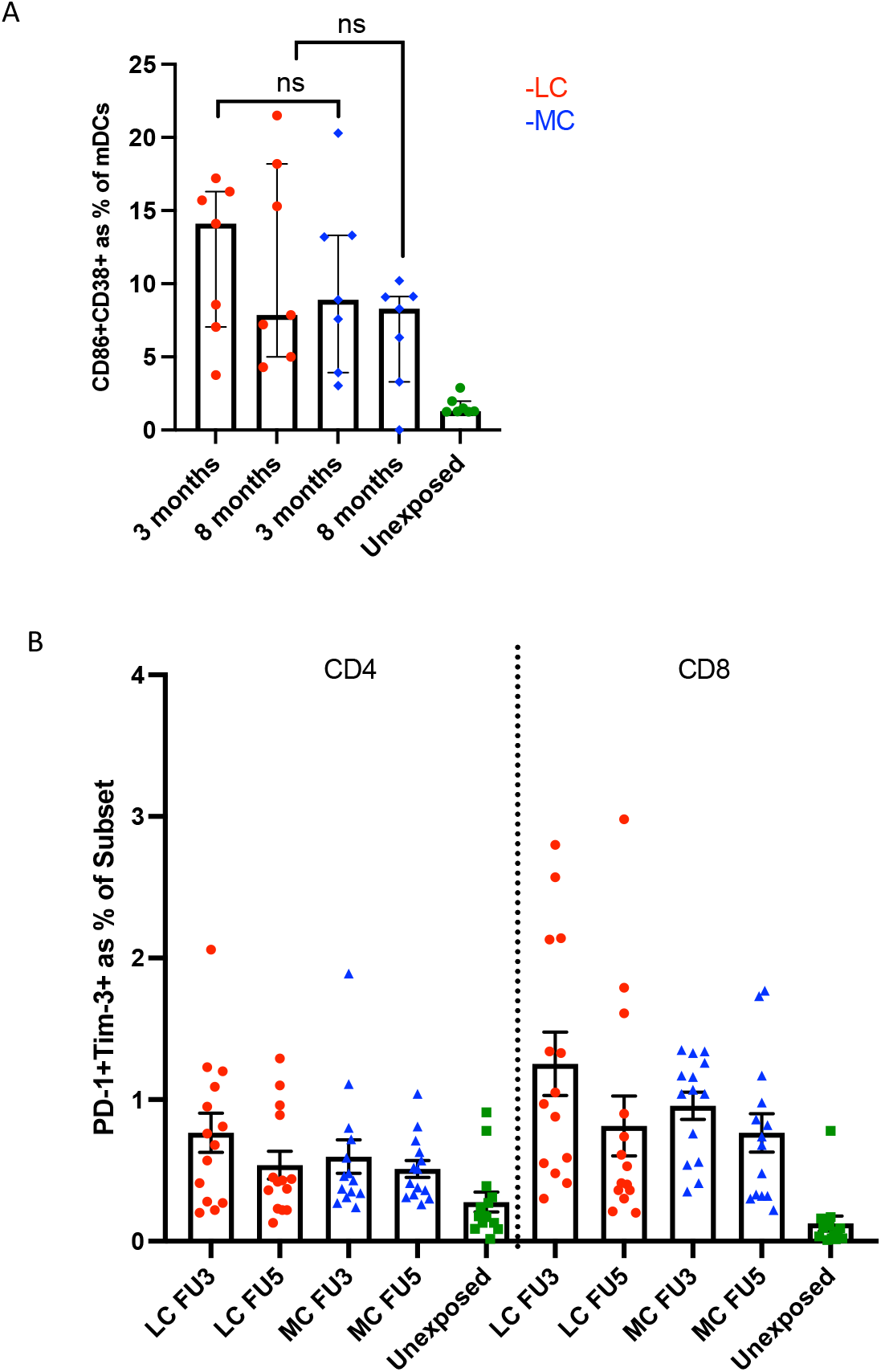
Activated phenotypes. (A) No difference in activated state of Myeloid dendritic cells (mDCs) at 3 months and 8 months. (B) No difference between LC and MC at 3 and 8 months when co-expression of PD-1 and Tim-3 were examined (long COVID, and MC (asymptomatic matched controls).

## Notes

### Competing Interest Statement

The authors have declared no competing interest.

### Clinical Trial

ACTRN12620000554965

### Funding Statement

We appreciate grant support from the St Vincents Clinic Foundation
- the Curran Foundation, the Rapid Response Research Fund (UNSW)
- the Medical Research Futures Fund (Australia) - NHMRC programme grant APP 1055214 (LMB) - Medical Research Future Fund award GNT 1175865 - - Austin Medical Research Foundation Grant
- the Victorian Government
- MRFF Award (2005544)
- NHMRC program grant (1149990)
- NHMRC Fellowships 1136322 and 1123673

### Author Declarations

- The ADAPT study was approved by the St Vincents Hospital Research Ethics Committee (2020/ETH00964) and is a registered trial (ACTRN12620000554965) - ADAPT-C sub study was approved by the same committee (2020/ETH01429) - All data were stored using REDCap electronic data capture tools - Unexposed healthy donors were recruited through St Vincents Hospital and was approved by St Vincents Hospital Research Ethics Committee (HREC/13/SVH/145) - The University of Melbourne unexposed donors were approved by Medicine and Dentistry HESC-Study ID 2056689. All participants gave written informed consent.

